# Genome-wide association study of febrile seizures identifies seven new loci implicating fever response and neuronal excitability genes

**DOI:** 10.1101/2020.11.18.20233916

**Authors:** Line Skotte, João Fadista, Jonas Bybjerg-Grauholm, Vivek Appadurai, Michael S Hildebrand, Thomas F Hansen, Karina Banasik, Jakob Grove, Clara A Climent, Frank Geller, Carmen F Bjurström, Bjarni J Vilhjálmsson, Matthew Coleman, John A Damiano, Rosemary Burgess, Ingrid E Scheffer, Ole Birger Vesterager Pedersen, Christian Erikstrup, David Westergaard, Kaspar René Nielsen, Erik Sørensen, Mie Topholm Bruun, Xueping Liu, Henrik Hjalgrim, Tune H Pers, Preben Bo Mortensen, Ole Mors, Merete Nordentoft, Julie W Dreier, Anders Børglum, Jakob Christensen, David M Hougaard, Alfonso Buil, Anders Hviid, Mads Melbye, Henrik Ullum, Samuel F Berkovic, Thomas Werge, Bjarke Feenstra

## Abstract

Febrile seizures represent the most common type of pathological brain activity in young children and are influenced by genetic, environmental, and developmental factors. While usually benign, in a minority of cases, febrile seizures precede later development of epilepsy. Here, we conducted a genome-wide association study of febrile seizures with 7,635 cases and 93,966 controls identifying and replicating seven new loci, all with *P* < 5 × 10^−10^. Variants at two loci were functionally related to altered expression of the fever response genes *PTGER3* and *IL10*, and four other loci harbored genes (*BSN, ERC2, GABRG2, HERC1*) influencing neuronal excitability by regulating neurotransmitter release and binding, vesicular transport or membrane trafficking at the synapse. *GABRG2* is a well-established epilepsy gene comprising variants associated with febrile seizures, and overall we found positive genetic correlations with epilepsies (*r*_*g*_ = 0.39, *P* = 1.68 × 10^−4^). Finally, a polygenic risk score based on all genome-wide significant loci was associated within patients with number of hospital admissions with febrile seizures and age at first admission, suggesting potential clinical utility of improved genetic understanding of febrile seizure genesis.

## INTRODUCTION

Fever is a systemic response to infection characterized by elevated core body temperature, orchestrated by a complex interplay between the immune system and the neuronal circuitry^1^. Although generally conferring a survival benefit, the fever response may occasionally induce adverse events. In young children, fever lowers the seizure threshold and 2-5% of children of European ancestries experience febrile seizures before 5 years of age^2^. Most febrile seizures are benign and self-limiting with no recurrence or further sequelae^3^. However, febrile seizures may herald the onset of various epilepsy syndromes, including very severe ones, with a long-term study indicating 7% of children with FS subsequently develop epilepsy^4^. Seizure disorders are also in varying degree associated with neurological and psychiatric comorbidities^5^.

The mechanisms by which fever can result in neuronal hyperexcitability and seizures are still incompletely understood. Increased brain temperature is known to directly affect the sensitivity of ion channels, which might be sufficient to generate febrile seizures^6,7^. Other suggested mechanisms include alkalosis from fever-induced hyperventilation^8^ and effects of inflammatory mediators such as cytokines on neuronal excitability^9^. Although the exact mechanisms underlying febrile seizures remain elusive, it is well recognized that the exposures that precipitate febrile seizures act in concert with developmental and genetic factors^10^.

Most genetic studies of febrile seizures have focused on rare epilepsy syndromes that include febrile seizures as part of the clinical presentation^11–14^. Less is known about the genetic background of common febrile seizures. We previously conducted a genome-wide association study (GWAS) of febrile seizures and identified 4 susceptibility loci for febrile seizures in general, implicating the sodium channel genes *SCN1A* and *SCN2A*, a TMEM16 transmembrane protein family gene (*ANO3*), and a region on 12q21.33 associated with magnesium levels^15^. We further identified 2 loci distinctly associated with febrile seizures as an adverse event following measles, mumps and rubella (MMR) vaccination, implicating the innate immune system genes *IFI44L* and *CD46*^15^. A few other loci have also been reported in candidate gene studies of smaller FS cohorts, including variants in the synaptic zinc transporter gene *ZNT3*^16^, and the immunoregulatory cytokine gene *IL6*^17^. Despite this progress, the identified variants only explain a small fraction of the variance in disease liability.

The present study is based on a strict GWAS meta-analysis design, including 7,635 febrile seizures cases and more than 90,000 controls. Here, our aims are to identify novel robustly associated genetic loci for febrile seizures, to investigate potential underlying genetic mechanisms, to analyze shared genetic susceptibility with epilepsies and neuropsychiatric diseases, and to study the association between polygenic risk score for febrile seizures and an individual’s number of hospital admissions with a febrile seizures diagnosis and the age of the first such admission.

## RESULTS

Our study design is illustrated in **Supplementary Fig. 1**. The discovery stage GWAS meta-analysis included 4,502 cases and 51,049 controls of European ancestries. Statistical power simulations suggested that our discovery analysis had >80% power to detect variants with odds ratios (ORs) down to 1.15-1.25 for common variants (**Supplementary Fig. 2**). After data cleaning and imputation based on the 1000 genomes project phase 3 reference panel^18^, 6.8 million autosomal variants were analyzed for association with febrile seizures. The genomic inflation factor was 1.08 and the linkage disequilibrium (LD) score regression intercept was 1.03, indicating minimal population stratification. Quantile-quantile and Manhattan plots are shown in **Figure 1**. Eight loci reached genome-wide significance (*P* < 5 × 10^−8^), including four previously described loci (*SCN1A, SCN2A, ANO3*, 12q21.33)^15^ and four novel loci (*PTGER3, IL10, BSN, ERC2*; **Fig. 1, Supplementary Fig. 3a-d**). Three additional novel loci (*GABRG2, MAP3K9, HERC1*) had multiple variants associated at 5 × 10^−8^ < *P* < 1 × 10^−6^ and were considered suggestive in the discovery stage (**Fig. 1, Supplementary Fig. 3e-g**). For each of the seven novel genome-wide significant or suggestive loci, we selected one single nucleotide polymorphism (SNP) for replication stage genotyping in 3,235 cases and 40,519 controls from Danish and Australian cohorts (**Supplementary Fig. 1**). Furthermore, we performed analyses conditioning on the lead SNP at each locus, but did not identify any additional SNPs fulfilling the criteria for replication genotyping (**Supplementary Fig. 4**). All seven novel genetic loci were replicated and reached genome-wide significance (*P* < 5 × 10^−8^) in the combined analysis (**Table 1, Supplementary Fig. 5**). There was no interaction between genotype and sex (**Supplementary Table 1**) and the additive genetic model fitted the data well (**Supplementary Table 2**).

**Table 1.** Discovery, replication and combined results for seven novel loci associated with febrile seizures

| Locus | Chr. | Position (bp) | SNP | Effect allele | Alt. allele | Gene | Sample set | EAF cases | EAF controls | OR (95% CI) | $P$ | $I^2$ | $P_{het}$ |
| --- | --- | --- | --- | --- | --- | --- | --- | --- | --- | --- | --- | --- | --- |
| 1p31.1 | 1 | 71163715 | rs3001038 | A | G | *PTGER3* | SSI | 0.527 | 0.486 | 1.19 (1.10-1.28) | 1.42E-05 |  |  |
|  |  |  |  |  |  |  | iPSYCH | 0.524 | 0.488 | 1.15 (1.09-1.22) | 8.59E-07 |  |  |
|  |  |  |  |  |  |  | Combined discovery |  |  | 1.17 (1.11-1.22) | 6.54E-11 | 0 | 0.58 |
|  |  |  |  |  |  |  | Replication |  |  | 1.10 (1.03-1.16) | 0.00214 | 37.8 | 0.2 |
|  |  |  |  |  |  |  | All combined |  |  | 1.14 (1.10-1.18) | **1.84E-12** | 58.9 | 0.12 |
| 1q32.1 | 1 | 206944645 | rs1518111 | T | C | *IL10* | SSI | 0.215 | 0.194 | 1.15 (1.04-1.26) | 0.00468 |  |  |
|  |  |  |  |  |  |  | iPSYCH | 0.212 | 0.186 | 1.18 (1.10-1.27) | 3.65E-06 |  |  |
|  |  |  |  |  |  |  | Combined discovery |  |  | 1.17 (1.11-1.24) | 4.62E-08 | 0 | 0.62 |
|  |  |  |  |  |  |  | Replication |  |  | 1.14 (1.06-1.23) | 0.000448 | 0 | 0.76 |
|  |  |  |  |  |  |  | All combined |  |  | 1.16 (1.11-1.21) | **1.02E-10** | 0 | 0.59 |
| 3p21.31 | 3 | 49644012 | rs11917431 | T | C | *BSN* | SSI | 0.308 | 0.282 | 1.13 (1.04-1.23) | 0.00284 |  |  |
|  |  |  |  |  |  |  | iPSYCH | 0.319 | 0.286 | 1.17 (1.10-1.25) | 4.6E-07 |  |  |
|  |  |  |  |  |  |  | Combined discovery |  |  | 1.16 (1.10-1.22) | 4.42E-09 | 0 | 0.53 |
|  |  |  |  |  |  |  | Replication |  |  | 1.13 (1.06-1.20) | 0.000235 | 0 | 0.95 |
|  |  |  |  |  |  |  | All combined |  |  | 1.15 (1.10-1.19) | **5.63E-12** | 0 | 0.52 |
| 3p14.3 | 3 | 56298138 | rs4974130 | C | T | *ERC2* | SSI | 0.076 | 0.058 | 1.34 (1.15-1.55) | 0.00019 |  |  |
|  |  |  |  |  |  |  | iPSYCH | 0.072 | 0.055 | 1.33 (1.19-1.48) | 1.5E-06 |  |  |
|  |  |  |  |  |  |  | Combined discovery |  |  | 1.33 (1.22-1.45) | 4.24E-10 | 0 | 0.95 |
|  |  |  |  |  |  |  | Replication |  |  | 1.27 (1.13-1.44) | 6.68E-05 | 21.1 | 0.28 |
|  |  |  |  |  |  |  | All combined |  |  | 1.31 (1.22-1.41) | **1.52E-13** | 0 | 0.57 |
| 5q34 | 5 | 161508503 | rs1997583 | T | A | *GABRG2* | SSI | 0.108 | 0.092 | 1.20 (1.06-1.36) | 0.00496 |  |  |
|  |  |  |  |  |  |  | iPSYCH | 0.101 | 0.085 | 1.24 (1.12-1.37) | 4.65E-05 |  |  |
|  |  |  |  |  |  |  | Combined discovery |  |  | 1.22 (1.13-1.32) | 5.11E-07 | 0 | 0.7 |
|  |  |  |  |  |  |  | Replication |  |  | 1.33 (1.21-1.46) | 8.8E-09 | 64.8 | 0.06 |
|  |  |  |  |  |  |  | All combined |  |  | 1.26 (1.19-1.34) | **5.48E-14** | 42.4 | 0.19 |
| 14q24.2 | 14 | 71323249 | rs244620 | C | T | *MAP3K9* | SSI | 0.469 | 0.447 | 1.10 (1.02-1.19) | 0.0151 |  |  |
|  |  |  |  |  |  |  | iPSYCH | 0.485 | 0.451 | 1.15 (1.08-1.22) | 2.09E-06 |  |  |
|  |  |  |  |  |  |  | Combined discovery |  |  | 1.13 (1.08-1.19) | 1.33E-07 | 0 | 0.42 |
|  |  |  |  |  |  |  | Replication |  |  | 1.11 (1.05-1.18) | 0.000384 | 0 | 0.54 |
|  |  |  |  |  |  |  | All combined |  |  | 1.13 (1.09-1.17) | **2.35E-10** | 0 | 0.68 |
| 15q22.31 | 15 | 64132818 | rs62023078 | G | T | *HERC1* | SSI | 0.848 | 0.823 | 1.21 (1.09-1.34) | 0.000346 |  |  |
|  |  |  |  |  |  |  | iPSYCH | 0.841 | 0.821 | 1.15 (1.06-1.24) | 0.000337 |  |  |
|  |  |  |  |  |  |  | Combined discovery |  |  | 1.17 (1.10-1.25) | 7.43E-07 | 0 | 0.46 |
|  |  |  |  |  |  |  | Replication |  |  | 1.17 (1.08-1.27) | 0.000161 | 55.8 | 0.1 |
|  |  |  |  |  |  |  | All combined |  |  | 1.17 (1.11-1.23) | **4.97E-10** | 0 | 0.99 |

Chr, chromosome; bp, base pair; EAF, effect allele frequency; OR, odds ratio; CI, confidence interval; $P$ , association $P$ value for febrile seizures; $I^2$ , heterogeneity estimate; $P_{het}$ , $P$ value from the Cochran Q test of heterogeneity.

**Figure 1.**
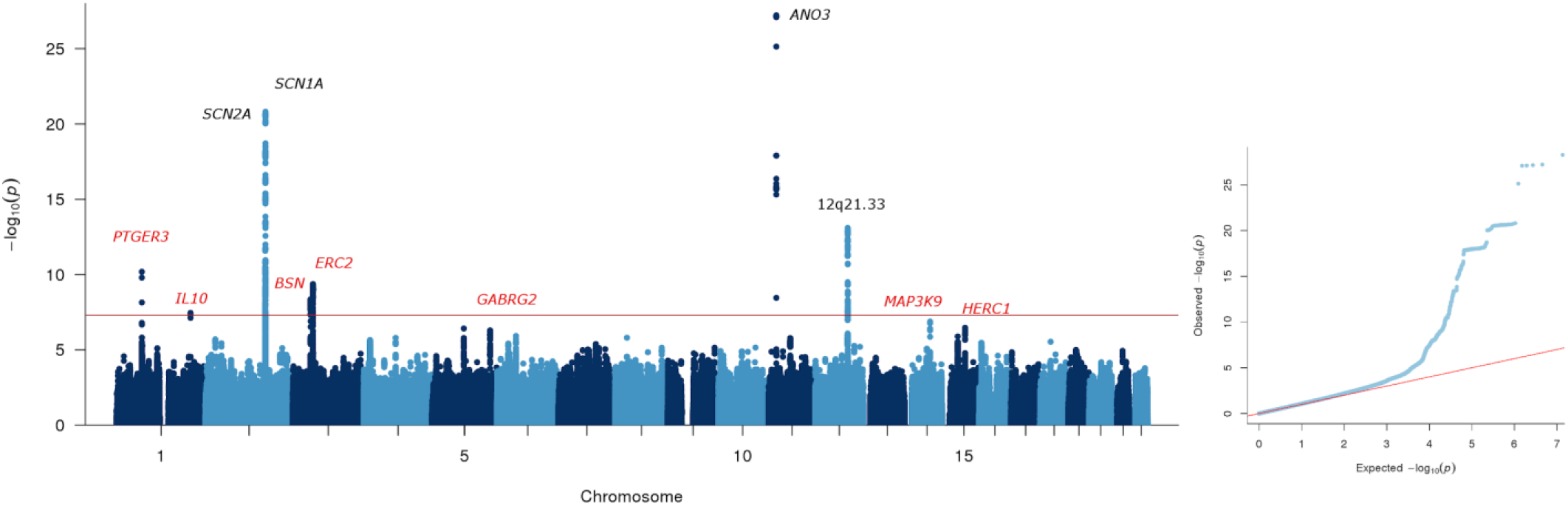
Febrile seizures genome-wide association analysis. Manhattan plot of −log_10_ *P* values across the chromosomes (left panel) and corresponding quantile-quantile plot of observed versus expected –log_10_ *P* values (right panel). Robustly associated loci are named; previously known loci are indicated in black; novel loci in red. The names above each locus represent the nearest protein coding gene at the locus or the chromosome band if the locus is in an intergenic region.

### Two novel loci harboring fever response genes

At the first locus for febrile seizures on chromosome 1p31.1, rs3001038 yielded the lowest *P* value (combined OR = 1.14, 95% CI = 1.10–1.18; *P* = 1.84 × 10^−12^). The lead variant rs3001038 was located in an LD block containing the important fever response gene *PTGER3*^1^ (**Supplementary Fig. 3a**). In search of possible functional mechanisms underlying the association signal, we annotated the 109 variants within 500 kb of the lead SNP that had discovery stage association *P* < 1 × 10^−4^ (**Supplementary Table 3**). Of note, many associated SNPs at the locus were reported as expression quantitative loci (eQTLs) for *PTGER3* in various brain tissues in the Genotype Tissue Expression (GTEx) project with risk-increasing alleles corresponding to increased *PTGER3* expression (**Supplementary Table 4**).

The association signal at the second locus on chromosome 1q32.1 was sharply defined by a few SNPs intronic or upstream of *IL10*, which encodes the antipyretic cytokine interleukin 10 (IL-10)^19^ (**Supplementary Fig. 3b**). The SNP rs1518111 was taken forward to the replication stage and confirmed (combined OR = 1.16, 95% CI = 1.11–1.21; *P* = 1.02 × 10^−10^). Of the six variants with *P* < 1 × 10^−4^, two have been reported to associate with the inflammatory disorder Behçet’s disease (*P* < 5 × 10^−8^), with the risk allele also corresponding to an increased risk of febrile seizures in our data (**Supplementary Table 5**). Notably, rs1518111 and other associated SNPs at the locus are all strong eQTLs for *IL10* in whole blood with the risk alleles for febrile seizures corresponding to lower *IL10* expression (**Supplementary Table 4**).

### Five additional novel loci including genes affecting neuronal excitability

At the third novel locus on chromosome 3p21.31, the SNP rs11917431, which is intronic in *BSN* yielded the lowest *P* value (combined OR = 1.15, 95% CI = 1.10–1.19; *P* = 5.63 × 10^−12^). *BSN* encodes Bassoon, a scaffolding protein of the presynaptic active zone^20^. The locus is characterized by extensive LD with SNPs highly correlated to the lead SNP spanning a region of almost 500 kb (**Supplementary Fig. 3c**). Fourteen of the SNPs at the locus with *P* < 1 × 10^−4^ were exonic, including 5 non-synonymous variants resulting in amino acid changes of USP4, GPX1, BSN, MST1, and RNF123, respectively (**Supplementary Table 3**). There were also a large number of previously reported GWAS associations at the locus, particularly for cognitive traits, chronic inflammatory diseases and blood protein levels (**Supplementary Table 5**). The risk alleles for febrile seizures SNPs at this locus were previously positively associated with math ability and other cognitive measures (**Supplementary Table 5**). A massive number of eQTL associations involving a wide array of genes and tissues have been reported for SNPs at the locus (**Supplementary Table 4**).

The lead SNP at the fourth locus on chromosome 3p14.3, rs4974130 (combined OR = 1.31, 95% CI = 1.22–1.41; *P* = 1.52 × 10^−13^) was intronic in *ERC2* with a swathe of highly correlated SNPs within the gene having similar *P* values (**Supplementary Fig. 3d**). *ERC2* encodes CAST, another protein of the cytomatrix of the active zone^21^. Four of these SNPs were exonic, including a missense variant in *ERC2* (**Supplementary Table 3**). Furthermore, the lead SNP and many other associated SNPs at the locus were reported as eQTLs for *ERC2* in cerebellum in the GTEx project with risk-increasing alleles corresponding to increased *ERC2* expression, and a few GWAS catalog associations were reported (**Supplementary Tables 4–5**).

At the fifth locus for febrile seizures on chromosome 5q34, the lead SNP, rs1997583 (combined OR = 1.26, 95% CI = 1.19–1.34; *P* = 5.48 × 10^−14^) was intronic in *GABRG2*, which encodes the γ2 subunit of gamma-aminobutyric type A (GABA_A_) receptor^22^ (**Supplementary Fig. 3e**), and where pathogenic missense variants cause febrile seizures in rare dominant families^11,12^. Other associated SNPs at the locus were located within or near the gene; one was exonic (synonymous) and none were reported as eQTLs or in the GWAS catalog (**Supplementary Tables 3–5**).

The sixth locus comprised associated SNPs in an LD block on chromosome 14q24.2 containing the gene *MAP3K9* encoding mitogen-activated kinase kinase kinase 9 (**Supplementary Fig. 3f**). The lead SNP, rs244620 (combined OR = 1.13, 95% CI = 1.09–1.17; *P* = 2.35 × 10^−10^) has been reported as an eQTL for *MAP3K9* in a range of different GTEx tissues (**Supplementary Table 4**) with the risk allele rs244620-C corresponding to increased expression in most tissues. Associated SNPs at the locus also showed strong cis-eQTL effects for *MAP3K9, TTC9*, and *MED6* in whole blood samples from the eQTLGen Consortium also with the febrile seizures risk alleles corresponding to increased expression (**Supplementary Table 4**).

Finally, at the seventh locus on chromosome 15q22.31, rs62023078 (combined OR = 1.17, 95% CI = 1.11–1.23; *P* = 4.97 × 10^−10^) was intronic in *HERC1*, which encodes HERC1, an E3 ubiquitin ligase located at the presynaptic terminal^23^ (**Supplementary Fig. 3g**). Among SNPs at the locus with *P* < 1 × 10^−4^, many have been reported as cis-eQTLs in whole blood and other tissues with febrile seizures risk alleles corresponding to decreased expression of *HERC1, USP3, DAPK2*, and *PLEKHO2*, and increased expression of *FBXL22, APH1B*, and *RBPMS2* (**Supplementary Table 4**).

### Exome analyses and fine mapping

Exome sequencing data were available in the Initiative for Integrative Psychiatric Research (iPSYCH) cohort for 589 febrile seizures cases and 10,378 controls. At each locus and for all genes within 1 Mb of the top variant, we used these data to conduct gene-based tests for aggregated effects of rare coding variants (optimal sequence kernel association test, SKAT-O; see Methods). Among the 159 genes tested, we were not able to detect any locus-wide significant gene-based rare-variant association to febrile seizures, except for an association for *SNX22* at the *HERC1* locus, which was however driven by only 0.36% of individuals carrying rare variants in the gene (**Supplementary Table 6**). Statistical fine mapping of the novel loci produced credible sets of variants (**Supplementary Table 7**), which together with the functional annotation in **Supplementary Tables 3–5** provide a basis for follow-up searches for causal variants at the loci.

### Previously reported loci

Four loci associated with febrile seizures in general (*SCN1A, SCN2A, ANO3*, and 12q21.33) were already reported based on the Statens Serum Institut (SSI) cohort included in this meta-analysis^15^. These were all corroborated by analysis of the independent iPSYCH cohort (**Supplementary Table 3**) and functional annotation revealed many entries in the GWAS Catalog as well as eQTLs for several genes, including the voltage gated Na^+^ channel genes *SCN1A* and *SCN2A* (**Supplementary Tables 4 and 5**). A comparison of effect sizes against minor allele frequencies of the 7 novel and 4 known genome-wide significant loci reflects theoretical expectations from simulations of statistical power based on sample sizes of the previous and the current GWAS (**Figure 2**). At a given minor allele frequency, the effects of the novel loci are smaller than those of the known loci, and further undetected loci are likely to have even smaller effects. An exception to this pattern may be loci for febrile seizures elicited by a specific exposure. In our previous study, we detected two loci (harboring the interferon-stimulated gene *IFI44L* and the measles virus receptor gene *CD46*) that were distinctly associated with febrile seizures occurring as a rare adverse event after measles, mumps and rubella (MMR) vaccination^15^. Vaccination data was not available for cohorts other than SSI, but the vast majority of febrile seizures episodes have no temporal connection to MMR vaccination^24^, and analysis of the *IFI44L* and *CD46* variants in the iPSYCH and Danish Blood Donor Study (DBDS) cohorts showed little evidence of association (**Supplementary Table 8**). Furthermore, none of the seven novel loci differed between MMR-related febrile seizures and MMR-unrelated febrile seizures (**Supplementary Table 9**).

**Figure 2.**
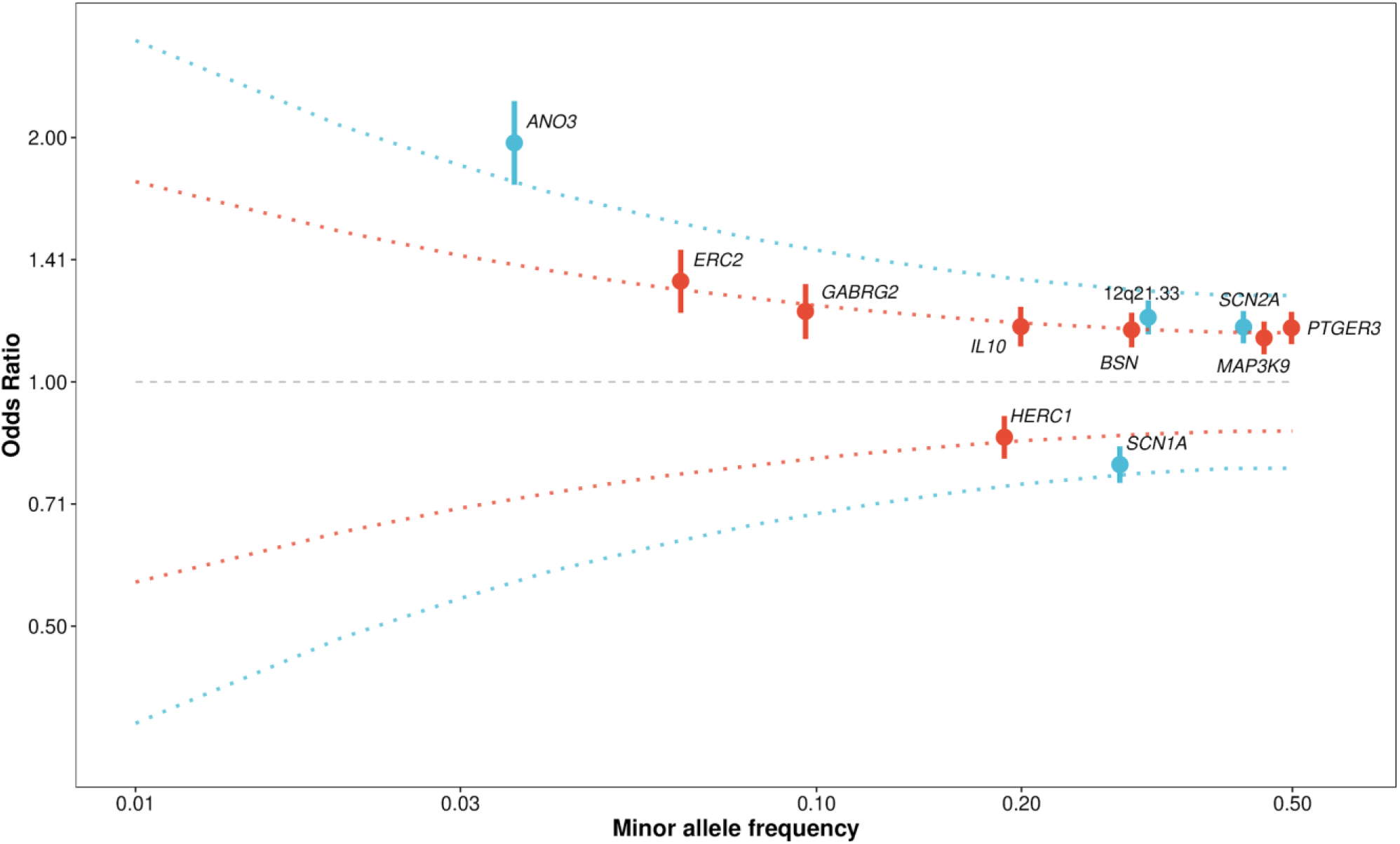
Minor allele frequencies and effect sizes (odds ratio, OR in discovery stage) for 11 robustly associated febrile seizures loci. Previously known loci are indicated in blue; novel loci in red. The red dotted lines represent ORs corresponding to 80% statistical power based on simulations with the sample size of the current study. The blue lines represent ORs needed for 80% power in simulations with the sample size of the previous febrile seizures GWAS^15^.

### Heritability and enrichment analyses

To assess the combined effects of loci, we used a multifactorial liability threshold model^25^ and found that the lead SNPs at the 11 robustly associated loci collectively explained 2.8% of the variance in liability to febrile seizures. Looking more broadly across the genome, the estimated proportion of variance in febrile seizures liability explained by all autosomal SNPs with a minor allele frequency >1% (SNP heritability) was 10.8% (SE 1.9%).

Based on a set of 131 genes encoding proteins of the presynaptic active zone^26^, we performed a gene-set analysis, but did not observe enrichment of febrile seizures associations (*P*_MAGMA_ = 0.17). We also analyzed enrichment of expression in particular tissue and cell types by testing whether genes in regions associated with febrile seizures were highly expressed in any of 209 Medical Subject Heading (MeSH) annotations based on gene expression data from 37,427 microarrays. While no annotations were significant after correction for the 209 analyzed categories, we note that the distribution of enrichment results was skewed towards brain tissues having the lowest *P* values (**Supplementary Fig. 6**).

### Shared susceptibility with epilepsy

Given the fact that rare variants in *GABRG2, SCN1A* and *SCN2A* at three of the febrile seizures loci have been linked to a range of epilepsy syndromes^27,28^ and the epidemiological links between febrile seizures and epilepsy, we wanted to examine potential shared genetic susceptibility. First, we performed genetic correlation analyses between our febrile seizures results and summary statistics from the latest GWAS mega-analysis from the International League Against Epilepsy (ILAE) Consortium on Complex Epilepsies^29^. These analyses revealed positive genetic correlations with focal epilepsy (*r*_*g*_ = 0.59, SE = 0.24, *P* = 0.01), genetic generalized epilepsy (*r*_*g*_ = 0.23, SE = 0.07, *P* = 0.002), and all epilepsies combined (*r*_*g*_ = 0.39, SE = 0.10, *P* = 1.68 × 10^−4^).

Next, we included an additional subset of the iPSYCH cohort of patients diagnosed with epilepsy and assessed the association of epilepsy to a polygenic risk score (PRS) for febrile seizures based on the SSI discovery cohort alone. One unit increase in the standardized PRS was associated with an increased risk of epilepsy (OR 1.07, 95% CI 1.02–1.12, *P* = 2.64 × 10^−3^; n_controls_ = 62,906, n_cases_ = 1,978), which remained significant when correcting for febrile seizures case status (OR 1.05, 95% CI 1.00–1.10, *P* = 0.045). Stratification of the data into individuals with and without a history of febrile seizures revealed no difference in effect of PRS on risk of epilepsy (*P* = 0.93).

### Neuropsychiatric outcomes

iPSYCH is designed as a case-cohort study and consists of a random population sample, and groups of patients with neuropsychiatric diagnoses, including attention deficit/hyperactivity disorder (ADHD), affective disorder, anorexia, autism spectrum disorder (ASD), bipolar disease and schizophrenia. We tested if the febrile seizure association of each lead variant in the seven novel and four known loci was constant across these different iPSYCH sub-groups and when correcting for the 11 loci tested, no statistically significant heterogeneity could be claimed (**Supplementary Fig. 7**). To generalize this analysis, we used a PRS based on the SSI discovery cohort alone and regressed the PRS on 14 factors defined by iPSYCH group and febrile seizures diagnosis. **Figure 3a** shows that the adjusted mean PRS for febrile seizures among iPSYCH individuals without febrile seizures was close to the overall adjusted mean PRS. For iPSYCH individuals with a febrile seizures diagnosis, the adjusted mean PRS was elevated in five out of seven groups. The largest effect was seen for the iPSYCH population representative sample with febrile seizures, which in pairwise comparisons had a higher mean adjusted PRS than all other groups except the group defined by anorexia and febrile seizures (**Figure 3b**).

**Figure 3.**
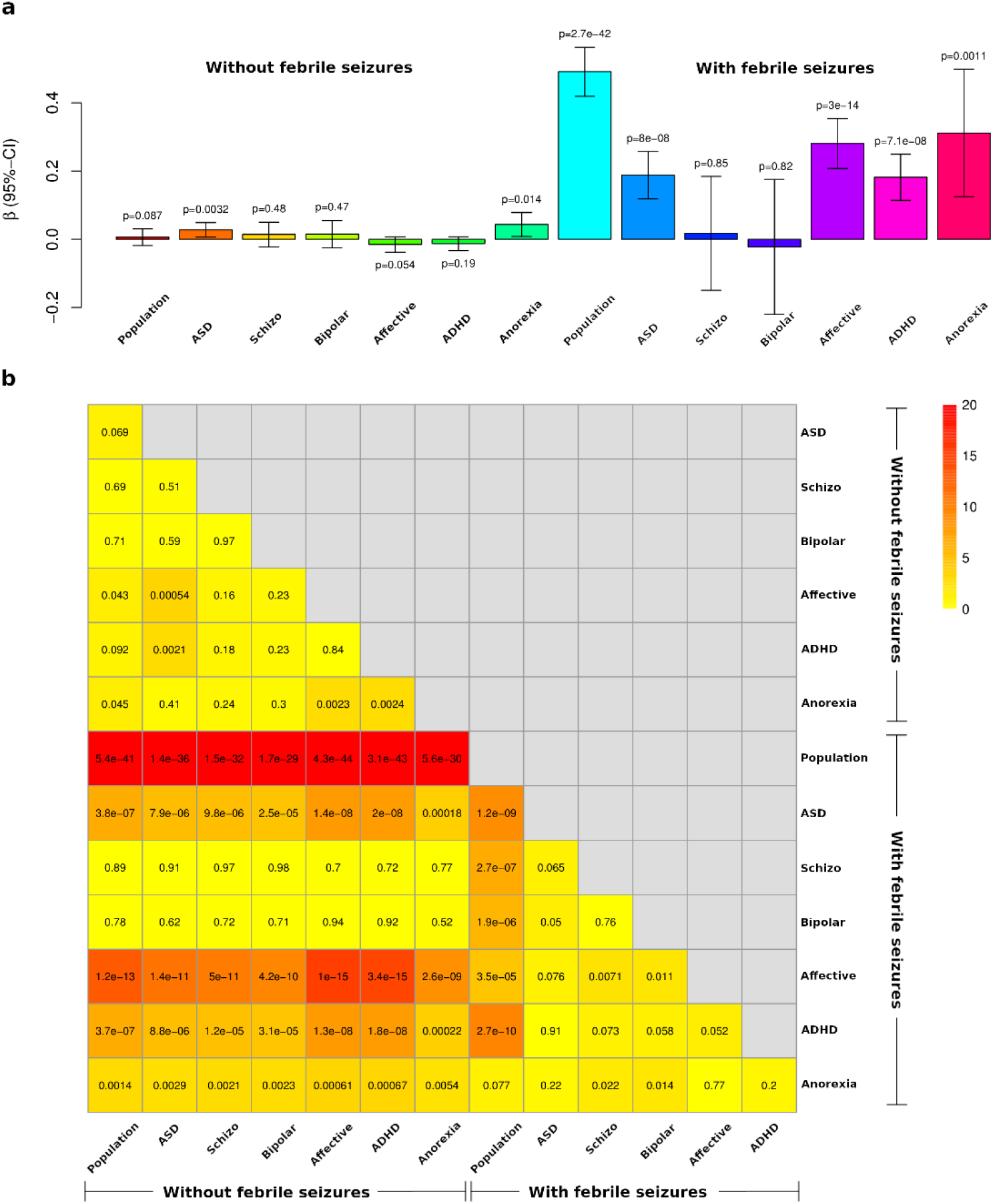
Multiple regression of PRS for febrile seizures on iPSYCH group and febrile seizures diagnosis. **a**, effect estimates of adjusted mean PRS for 14 factors defined by iPSYCH group and febrile seizures diagnosis with 95% confidence intervals. *P* values are from tests of difference from overall mean adjusted PRS. **b**, *P* values from Wald test of equal group effect in pairwise comparisons of the 14 groups.

### Analysis of hospitalization data

Next, we assessed the combined impact of the identified febrile seizures loci on 1) number of hospital admissions due to febrile seizures, and 2) the age at first admission with febrile seizures.

We constructed a PRS for febrile seizures based on the 11 robustly associated loci and analyzed association between the PRS and the number of additional febrile seizures in cases using negative binomial regression (see Methods). In the iPSYCH discovery cohort each unit increase in standardized PRS was associated with a 1.17 fold increase in expected additional febrile seizures hospital admissions (negative binomial test, *P* = 6.47 × 10^−4^; *n* = 2511 cases). This analysis could not be done in the SSI discovery cohort, due to the cohort’s special ascertainment scheme, but a similar estimate (1.16 fold increase in expected additional febrile seizures per PRS unit) was found in the DBDS replication cohort (negative binomial test, *P* = 0.07; *n* = 934 cases). The distribution of number of febrile seizures hospital admissions among cases and PRS by number of hospital admissions is seen in **Figure 4**, which also shows that febrile seizures patients with a high polygenic risk were more likely to be hospitalized with febrile seizures at a younger age.

**Figure 4.**
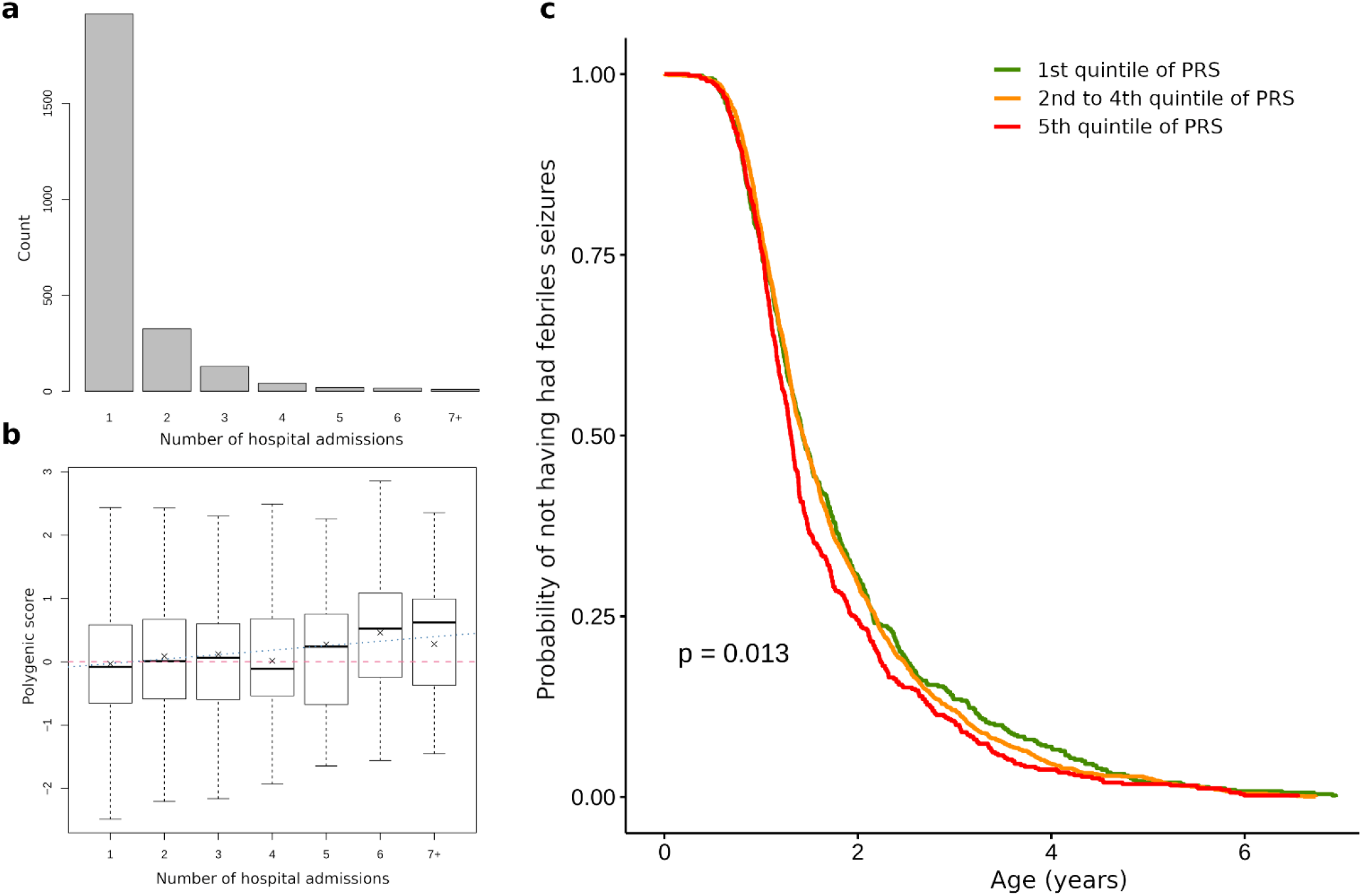
Association between PRS for febrile seizures and history of hospital admissions with a febrile seizures diagnosis among 2,511 febrile seizures cases in the iPSYCH cohort. **a**, number of separate admissions to hospital with a febrile seizures diagnosis among the 2,511 cases. **b**, box plots of PRS for febrile seizures based on 11 genome-wide significant loci by number of febrile seizures hospital admissions. **c**, Kaplan-Meier curves of time-to-event analysis for first febrile seizures hospital admission by groupings of PRS. A log-rank test was used to test for difference between the three curves.

## DISCUSSION

In this genome-wide meta-analysis of febrile seizures including a total of 7,635 cases and more than 90,000 controls, we identified seven novel robustly associated loci and confirmed four previously reported loci. Our results implicate central fever response genes as well as loci related to ion channel function, neurotransmitter release and binding, and vesicular transport and membrane trafficking at the synapse. We find positive genetic correlations between febrile seizures and epilepsy syndromes, and we show that higher polygenic risk scores for febrile seizures are associated with a higher number of hospital admissions for febrile seizures and lower age at first admission.

Febrile seizures are an outcome of intricate etiology involving feedback loops between the body’s fever response and the neuronal circuitry. It has been suggested that the regulation of body temperature may be different in children susceptible to febrile seizures^30,31^. Our association findings at the *PTGER3* and *IL10* loci are consistent with this hypothesis. *PTGER3* encodes EP3, one of four receptors for prostaglandin E2 (PGE2), which is considered to be a major pyrogenic mediator of fever^1^. It has been shown that mice lacking the EP3 receptor fail to show a febrile response to direct stimulation with PGE2 and to exposure to endogenous (IL-1β) and exogenous (lipopolysaccharide, LPS) pyrogens^32^. Additional experiments established that selective disruption of *Ptger3* in the median preoptic nucleus of the mouse brain was sufficient to block fever in response to PGE2 and LPS^33^. While the median preoptic nucleus is not assayed in GTEx, we found that risk alleles for the SNPs at the locus corresponded to higher expression of *PTGER3* in several other brain regions. We hypothesize that the genetic up-regulation of *PTGER3* leads to a more pronounced fever response, which in turn increases the susceptibility to febrile seizures. Furthermore, we found that febrile seizure risk alleles for SNPs at the *IL10* locus corresponded to lower IL10 expression in blood. IL10 encodes the anti-inflammatory cytokine IL-10, which in the complex landscape of cytokine signaling acts as a central endogenous antipyretic^19^. Thus, the genetic down-regulation of IL10 could conceivably lead to a more pronounced fever response and thereby confer increased risk of febrile seizures.

Other novel loci harbored genes such as *BSN, ERC2*, and *HERC1*, which have functions related to neurotransmitter release, vesicular transport and membrane trafficking. Presynaptic plasma membranes contain specialized regions called active zones (AZ), where synaptic vesicles release neurotransmitters such as, glutamate or GABA, depending on the type of neuron^20^. The release sites are characterized by a dense cytomatrix at the active zone (CAZ) organized by a number of proteins, including the large scaffolding protein Bassoon encoded by *BSN*. Disruption of Bassoon function in mice is associated with pronounced spontaneous seizures, and is used as an experimental model for epilepsy^34,35^. Bassoon interacts with multiple other CAZ proteins, including CAST (also known as ELKS2) encoded by *ERC2*^21^. Uncovering the complex roles of CAST, Bassoon and other proteins in the assembly of the CAZ and in synaptic transmission in different types of neurons is an active field of research^20,21,36^. Posttranslational modifications, such as ubiquitination add to the complexity of presynaptic dynamics, and recent mouse studies show the importance of the HERC1 E3 ubiquitin ligase in the regulation of vesicular transport and membrane trafficking at the synapse^23,37^. While these observations suggest *BSN, ERC2*, and *HERC1* are potential candidates at the loci on chromosomes 3p21.31, 3p14.3, and 15q22.31, follow-up studies are required to establish causal links and to illuminate mechanisms related to febrile seizures.

Our findings reinforce the notion that genetic studies of isolated febrile seizures can be informative for understanding epilepsy and seizure genesis in general. Previously, rare familial missense mutations in *GABRG2* have been found to segregate with febrile seizures and childhood absence epilepsy or genetic epilepsy with febrile seizures plus (GEFS+)^11,12^, and nonsense mutations of the gene have been found in more severe epilepsy phenotypes^28^. GABA is the major inhibitory neurotransmitter acting at GABA_A_ receptors, responsible for fast synaptic inhibition in the brain^22^, and it is thought that the variability of epilepsy phenotypes associated with *GABRG2* mutations is related to the extent of GABA signaling impairment^28^. Extending this line of thinking, we hypothesize that the common *GABRG2* variants identified here increase febrile seizures risk through subtle effects on GABA signaling. We also confirmed common variant associations for febrile seizures in the well-known epilepsy genes *SCN1A* and *SCN2A*. Previous studies have linked rare missense mutations in these voltage gated Na^+^ channel genes to a range of epilepsy syndromes, some involving febrile seizures^27^. Looking across the genome, we found positive genetic correlations between febrile seizures and focal epilepsy, genetic generalized epilepsy and all epilepsies combined. Delineating genetic differences and similarities between febrile seizures and epilepsies is an important area for future research.

From a clinical perspective, an enhanced understanding of potential etiological relations between febrile seizures and epilepsies is highly warranted. A central unresolved question is whether febrile seizures may in some cases lead to hippocampal damage, provoking a chronic process of epileptogenesis or whether shared genetic and developmental factors increase the susceptibility to febrile seizures as well as epilepsy^10,38^. Population registers and large biobanks, such as those available in Denmark, offer opportunities for carefully designed massive-scale studies addressing this question through genetic analyses of febrile seizures and epilepsy patients with long-term follow-up. Such studies may also enable prediction of individuals at high risk of febrile seizures and/or epilepsy, and possibly suggest preventive strategies. While the current study was not sufficiently powered to develop prediction tools, the fact that a score based on all genome-wide significant loci was associated with number of hospital admissions with febrile seizures and age at first admission, suggests future potential clinical utility of a deeper understanding of the genetic underpinnings of febrile seizures.

Epidemiological studies have reported associations between epilepsy, and to a lesser degree febrile seizures, and risk of psychiatric disorders^5,39–41^. However, somewhat surprisingly, an adjusted PRS for febrile seizures based on the SSI discovery cohort was on average lower in all iPSYCH disease groups except anorexia, compared to the iPSYCH population representative sample. While this question should be revisited in a later study with a larger sample for PRS construction, it raises the possibility that shared environmental or developmental risk factors may underlie epidemiological associations between febrile seizures and psychiatric disorders rather than shared common genetic variation. Interestingly, a large-scale genetic correlation study also found limited sharing of common genetic variants between epilepsy and psychiatric disorders, despite strong epidemiological associations^42^.

Our study had limitations, including the restriction to individuals of European ancestries. Incidence rates of febrile seizures vary considerably between different countries of the world^2^ and further studies are warranted to characterize genetic susceptibility to febrile seizures in diverse populations. A second limitation was the granularity of the outcome definition. With a positive predictive value of a diagnosis of febrile seizures (ICD-8 and ICD-10) of 93%, the Danish National Patient Register is an excellent resource for research^43^, but the register does not contain information about complex febrile seizures including characteristics such as the duration and type (focal or generalized) of febrile seizures, postictal neurological abnormalities and possible recurrence within 24 hours. Further studies are required to understand effects of variants at the identified loci across a phenotypic spectrum of thoroughly characterized febrile seizure patients. Information about the specific infectious exposure or inflammation leading to fever and febrile seizures would also be revealing, but is usually difficult to ascertain.

In conclusion, this largest genetic investigation of febrile seizures to date implicates central fever response genes as well as genes affecting neuronal excitability, including several known epilepsy genes. This adds to knowledge from clinical, epidemiological and laboratory studies that febrile seizures are the manifestation of a highly complex physiological process involving environmental triggers, neurodevelopmental vulnerability and genetic susceptibility. Furthermore, the positive genetic correlation between febrile seizures and both focal and generalized epilepsies underscores that carefully conducted genetic studies of febrile seizures can yield fundamental insights into the genetic regulation of neuronal excitability highly relevant to our understanding of epileptogenesis. Clearly, additional functional and genetic studies, including of rare as well as common variants at the new loci we have identified, are required to enhance our understanding of the pathophysiological pathways and triggers of seizures with and without fever.

### URLs

1000 Genomes Project, http://www.1000genomes.org/; ANNOVAR, http://annovar.openbioinformatics.org/; Braineac, http://www.braineac.org/; DEPICT, https://data.broadinstitute.org/mpg/depict/; DICE, https://dice-database.org/; eQTL Catalogue, https://www.ebi.ac.uk/eqtl/; eQTLGen Consortium, https://www.eqtlgen.org/; FUMA, https://fuma.ctglab.nl/; GTEx, https://www.gtexportal.org/; iPSYCH, http://ipsych.au.dk/about-ipsych/; LD score regression, https://github.com/bulik/ldsc/; MAGMA, https://ctg.cncr.nl/software/magma/; METAL, http://www.sph.umich.edu/csg/abecasis/metal/; Michigan Imputation Server, https://imputationserver.sph.umich.edu; NHGRI-EBI GWAS Catalog, https://www.ebi.ac.uk/gwas/; R software, http://www.r-project.org/; SNPTEST, https://mathgen.stats.ox.ac.uk/genetics_software/snptest/snptest.html.

### Data availability

GWAS summary statistics from this study will be made available via the Danish National Biobank website (https://www.danishnationalbiobank.com/gwas) upon publication of the study.

## Data Availability

GWAS summary statistics from this study will be made available via the Danish National Biobank website (https://www.danishnationalbiobank.com/gwas) upon publication of the study in a peer-reviewed journal.

## ONLINE METHODS

### Subjects

The discovery stage included two cohorts of Danish ancestry with existing GWAS data and phenotypic information from the Danish National Patient Register^44^. The register includes information from all inpatient admissions in Denmark since 1977 and all emergency and outpatient hospital contacts since 1995. Diagnoses are coded according to the International Classification of Diseases (ICD) using the 8th revision (ICD-8) from 1977 through 1993 and the 10th revision (ICD-10) from 1994 on. Eligible febrile seizures cases were defined as children registered with ICD-8 code 78021 or ICD-10 code R560 before the age of 7, and controls were required not to have any febrile seizures or epilepsy diagnosis codes (ICD-8 codes 345, 78020, 78029 and ICD-10 codes G40-G41, or R568) in the National Patient Register. The first discovery cohort included 1,991 febrile seizures cases and 4,097 controls from our previous GWAS of febrile seizures conducted at Statens Serum Institut (SSI)^15^. This study had a focus on febrile seizures occurring as an adverse event following measles, mumps and rubella (MMR) vaccination, and 925 of the cases included in the current study had a febrile seizures episode that occurred in a risk window of 9 to 14 d after the date of MMR vaccination. These are referred to as MMR-related febrile seizure cases. The remaining 1066 cases included in the current study experienced febrile seizures with no temporal relation to MMR vaccination^15^. The second discovery cohort was comprised of 2,511 febrile seizures cases and 46,952 controls from the Initiative for Integrative Psychiatric Research (iPSYCH) study^45^. The iPSYCH sample included patient groups of six neuropsychiatric disorders (autism spectrum disorder (ASD), attention deficit/hyperactivity disorder (ADHD), schizophrenia, affective disorder, bipolar disease, and anorexia) and a random population sample.

The replication stage included three cohorts with genome-wide imputed data available or custom genotyping. First, from SSI, we included an additional 2,004 cases (411 MMR-related febrile seizures and 1,593 MMR-unrelated) and 3,208 controls. Second, from the Danish Blood Donor Study (DBDS), we analyzed 928 cases and 28,862 controls. Third, we included an Australian cohort of 201 febrile seizures cases and 847 controls. In the Danish replication cohorts, case and control definitions were the same as in the discovery stage. In the Australian cohort, a comprehensive assessment of each patient for diagnosis of febrile seizure was obtained using a validated seizure questionnaire, clinical evaluation by an experienced epileptologist, and review of relevant medical records and clinical investigations as described previously^16^. Cases were unrelated patients of European ancestries recruited from Australia diagnosed with febrile seizures and controls were individuals without febrile seizures from the same population, a proportion of which were reported in a prior study^16^. The study design is schematically illustrated in **Supplementary Fig. 1**.

### Ethics statement

The SSI study was approved by the Scientific Ethics Committee for the Capital City Region (Copenhagen) and the Danish Data Protection Agency. The Scientific Ethics Committee also granted exemption from obtaining informed consent from participants (H-3-2010-003) as the study was based on biobank material. The Danish Scientific Ethics Committee, the Danish Data Protection Agency and the Danish Neonatal Screening Biobank Steering Committee approved the iPSYCH study. DBDS was approved by the Central Denmark Regional Committee on Health Research Ethics (M-20090237) and the Danish Data Protection Agency, Copenhagen (2012-58-0004, RH-30-0444 / I-suite no.: 00922).

For the Australian replication cohort, all experimental protocols were approved by the Human Research Ethics Committee of Austin Health, Melbourne, Australia. All methods were carried out in accordance with the approved guidelines of the Human Research Ethics Committee of Austin Health, Melbourne, Australia. Informed consent was obtained from all human subjects.

### Genotyping, data cleaning and imputation

The SSI discovery cohort had existing GWAS data available from genotyping with Illumina Omni Bead Arrays and GenomeStudio software^15^. Data cleaning included filtering out individuals that (i) had more than 3% missing genotypes, (ii) had an autosomal heterozygosity rate deviating by more than 2.5 s.d. from the mean, (iii) had discordant sex information or (iv) were more than 6 s.d. away from the mean of any of the first five principal components in a principal-component analysis. We also estimated pairwise identity by descent (IBD) based on the GWAS data for all possible pairs of individuals. We then excluded one individual from each pair with an estimated IBD proportion >0.1875, corresponding to the point halfway between second-degree and third-degree relatives. Next, we excluded SNPs on the basis of a missing rate of >2%, a MAF of <0.01, deviations from Hardy-Weinberg equilibrium (*P* < 1 × 10^−6^), and genotyping batch differences in call rate or allele frequencies. The remaining 540,064 SNPs were used for imputation, which was done remotely at the Michigan Imputation Server (see URLs) using the 1000 Genomes Project Phase 3 reference panel. After imputation, 8,012,441 autosomal variants with MAF≥0.01 and imputation INFO >0.8 were available for analysis in 1,991 cases and 4,097 controls.

The iPSYCH discovery cohort comprised 78,050 individuals with existing GWAS data available from genotyping with the Illumina Infinium PsychChip v1.0^45^. Data cleaning involved filtering out SNPs that had a MAF of <0.01 or deviated from Hardy-Weinberg equilibrium (*P* < 1 × 10^−6^). Structural variants, tri-allelic SNPs, non-autosomal SNPs, and SNPs that were strand ambiguous (A/T or C/G) or did not uniquely align to the genome were also excluded, resulting in an imputation backbone of 246,369 autosomal SNPs. A total of 364 samples failing basic QC (discordant sex information, more than 1% missing genotypes, abnormal heterozygosity, duplicate sample discordance) were also excluded. Based on the good quality SNPs, all individuals were phased in a single batch using SHAPEIT3^46^ and imputed in 10 batches using Impute2^47^ with reference haplotypes from the 1000 genomes project phase 3. Prior to analysis, we filtered out participants who were (1) of non-European ancestries, (2) related to another participant in the sample (corresponding to an IBD proportion >0.1875), (3) were included in the SSI discovery or replication sample, or (4) had an epilepsy or non-febrile seizure diagnosis. We also balanced the ratio of febrile seizures cases to controls, so that it was the same within each of the participant groups of the iPSYCH study as illustrated in **Supplementary Fig. 8**. After these steps, 7,264,301 autosomal variants with MAF≥0.01 and imputation INFO >0.8 were available for analysis in 2,511 febrile seizures cases and 46,952 controls.

### GWAS meta-analysis

In each discovery cohort, the data were analyzed by logistic regression under an additive genetic model, including sex and 4 principal components as covariates. Variant positions were based on GRCh37/hg19 and alleles were labeled on the positive strand of the reference genome. A total of 6,805,333 variants were available in both discovery cohorts and were included for further analysis. We carried out combined analysis of the two discovery cohorts using fixed-effects inverse variance weighted meta-analysis as implemented in METAL^48^. Heterogeneity between studies was assessed using the I^2^ statistic and Cochrane’s Q test^49^. To assess possible confounding effects on the distribution of test statistics, we calculated the genomic inflation factor^50^ and the LD score regression intercept^51^. Loci with either one or more variants associated at *P* < 5 × 10^−8^ or two or more variants associated at 5 × 10^−8^ < *P* < 1 × 10^−6^ were identified and one top SNP from each locus was taken forward to the replication stage. To search for additional independent associated variants, we conditioned on the imputed allelic dosage of the top SNP at each locus in each discovery stage cohort and meta-analyzed the results. Any SNPs reaching *P* < 1 × 10^−6^ in these analyses were also considered eligible for the replication stage.

### Replication

For the SSI replication cohort, genotyping of the selected SNPs was performed using competitive allele-specific PCR (KASP) chemistry (LGC Genomics). In the Australian cohort, genomic DNA was extracted from whole blood (Qiagen QIAamp DNA Maxi Kit, Valencia, CA) and selected SNPs were genotyped by PCR amplification (Veriti Thermal Cucler, Applied Biosystems, Carlsbad, CA) and bidirectional sanger sequencing (BigDyeTM v3.1 Terminator Cycle Sequencing Kit and 3730xl DNA Analyzer, Applied Biosystems). The DBDS replication cohort had existing GWAS data available from genotyping with Illumina Global Screening Array and subsequent imputation, as described previously^52^. Association testing was done by logistic regression under an additive genetic model adjusting for sex. With the availability of GWAS data, the replication results from DBDS were also adjusted for the first 4 principal components. Combined analysis of the discovery and replication stage data was done by fixed-effects inverse variance-weighted meta-analysis. For the 1p31.1 locus, rs3001038 failed in genotyping of the Australian cohort and rs998550 was used as a proxy instead. For the 15q22.31 locus, rs62023078 failed in the replication cohorts from Statens Serum Institut and Australia, and rs13379670 was used as a proxy instead. We considered SNPs with *P* < 0.05 in the replication stage and *P* < 5 × 10^−8^ in the combined analysis to indicate robust evidence of association.

### Power analysis

We assessed the statistical power of our study by computer simulations in R^53^. Disease state was simulated from a logistic regression model allowing the odds ratio for a log-additive genetic effect and the frequency of the effect allele to vary and assuming a 3.6% population incidence of febrile seizures before the age of 5 years^54^. For each combination of effect size and effect allele frequency, we simulated 1000 data sets using the study sample size (4,502 cases and 51,049 controls). We then conducted association tests on the simulated data sets and calculated power as the proportion of tests with a *P* value lower than 5 × 10^−8^. To compare with statistical power in the previous GWAS of febrile seizures, we also performed simulations using the sample size (1,999 cases and 4,118 controls) of that study^15^.

### Functional annotation

For each of the 7 novel and 4 previously known genome-wide significant loci, we annotated all variants with discovery stage association *P* < 1 × 10^−4^ within 500 kb of the lead SNP at the locus using ANNOVAR^55^. Gene-based annotation was performed with gene definitions from the NCBI RefSeq database^56^, and SIFT^57^, PolyPhen-2^58^, and FATHMM^59^ were used to predict the pathogenicity of missense mutations. Using the FUMA web application^60^, we searched the GWAS Catalog for previously reported associations with P < 5 × 10^−8^ for SNPs at the febrile seizures loci. We also used FUMA to perform eQTL annotation based on the BRAINEAC, DICE, eQTL Catalogue, eQTLGen, and GTEx/v8 databases (see URLs).

### Exome analyses and fine mapping

Exome sequencing data were available for a subset of samples in the iPSYCH cohort. In total, n = 10,967 individuals, sampled from either ADHD (n = 2382), ASD (n = 3612), affective disorder (n = 1), bipolar disease (n = 652), schizophrenia (n = 1471), or the population representative sample (n = 2849) were analyzed. Out of these, 589 were febrile seizures cases. There were more potential febrile seizures controls with exome data available, but within each iPSYCH subset the ratio of febrile seizures cases to controls was approximately balanced at 1:17.6 to avoid spurious associations driven by the iPSYCH diseases. For these samples, and for each of the novel febrile seizures loci, we considered all genes within a 1 MB region of the lead variant and performed gene-based tests for rare-variant association, using the optimal sequence kernel association test (SKAT-O)^61^ approach as implemented in EPACTS version 3.2.6, using default settings.

We also used a Bayesian fine-mapping approach implemented in FINEMAP^62^ version 1.4 to construct 95% credible sets (containing the minimal set of variants with a total posterior probability of being causal of 95% or more) for each of the novel loci.

### Variance explained and genetic correlation analyses

Assuming a multifactorial liability threshold model, we used risk allele frequencies and estimated ORs from the discovery meta-analysis for the lead SNPs at all 11 genome-wide significant loci to estimate the proportion of variance in the liability to febrile seizures explained by these variants^25^. The estimate of variance explained by all common (MAF >1%) autosomal SNPs (also known as SNP heritability) was calculated based on the discovery stage meta-analysis results using LD score regression^51^. In the calculations of variance explained, we assumed a 3.6% population incidence of febrile seizures before the age of 5 years^54^.

We retrieved GWAS summary statistics for focal epilepsy, genetic generalized epilepsy, and all epilepsies combined from the ILAE Consortium on Complex Epilepsies^29^ and used LD score regression to estimate genetic correlations between febrile seizures and epilepsies.

### Enrichment analyses

To address the hypothesis that febrile seizures associations are enriched in genes that encode proteins of the presynaptic active zone, we defined a set of such genes based on experimental work in a rodent model^26^. We then performed gene-set enrichment analysis with MAGMA^63^ using the SNP-wise mean model.

We used DEPICT^64^ to first determine a set of uncorrelated variants with *P* < 1 × 10^−5^ by LD clumping (r^2^ = 0.1, window size = 500 kb). Using this set, we performed tissue and cell type enrichment analysis by testing whether the genes in associated regions were highly expressed in any of 209 Medical Subject Heading (MeSH) annotations for 37,427 microarrays on the Affymetrix U133 Plus 2.0 Array platform.

### Polygenic risk score analyses

To examine associations between general susceptibility to febrile seizures with other outcomes of interest, we constructed polygenic risk scores (PRS) by summing over risk allele counts/dosages at sets of relevant SNPs, with weights for each SNP corresponding to its estimated effect size. Two sets of scores were constructed. First, we included only the lead variants at each of the 7 novel and 4 known genome-wide significant and robustly replicated loci. Second, we constructed scores based on the GWAS summary statistics from the SSI discovery cohort. We generated PRS applying an LD-clumping and *P* value thresholding approach with 9 different *P* value thresholds (in -log_10_ scale: 0, 0.5, 1, 1.5, 2, 2.5, 3, 3.5, 4) as well as LDpred^65^ infinitesimal model and LDpred point-normal mixture with 7 different proportions of causal variants (1, 0.3, 0.1, 0.03, 0.01, 0.003, 0.001), yielding a total of 17 sets of PRSs. We then performed out-of-sample validation of how well each set of PRS predicted febrile seizures in the iPSYCH discovery cohort. The best predictive performance (assessed with Nagelkerke *R*^*2*^ measure) was found for LDpred with a proportion of 0.3% causal variants in the model.

We used logistic regression models to assess the association between PRS for febrile seizures and risk of epilepsy in the iPSYCH cohort with and without stratification by febrile seizures diagnosis. To investigate relations between iPSYCH disease groups and genetic susceptibility to febrile seizures, we used a multiple regression model with febrile seizures PRS as the outcome and 14 factors defined by iPSYCH group (ADHD, ASD, schizophrenia, affective disorder, bipolar disease, anorexia, and the population representative sample) and febrile seizures diagnosis (yes/no), and also adjusting for 10 principal components. We used Wald tests for pairwise comparisons of the 14 different groups’ effects on the PRS. For the hospitalization data for febrile seizures patients, we used a negative binomial model to analyze the effect of febrile seizures PRS on the number of febrile seizures hospital admissions, including sex and four principal components as covariates in the model. We also divided febrile seizures patients into three groups by PRS, corresponding to the first quintile, the second to fourth quintile, and the fifth quintile, and conducted time-to-event analysis of age at first febrile seizures hospital admission by PRS group by the Kaplan-Meier method. We used a log-rank test to test for differences between the three PRS groups.

## ACKNOWLEDGMENTS

The study was supported by grants from the Danish Medical Research Council (0602-01818B), the Oak Foundation (OCAY-18-598), the US National Institutes of Health (NIH)/National Institute of Allergy and Infectious Diseases (R01AI093697), the Novo Nordisk Foundation Challenge programme (NNF17OC0027594), a Lundbeck Foundation Ascending Investigator grant (R313-2019-554) to B.F., and National Health and Medical Research Council (NHMRC) Program Grant (1091593) to S.F.B. and I.E.S, Practitioner Fellowship (1006110) to I.E.S., and R.D Wright Career Development Fellowship (1063799) to M.S.H., and a Novo Nordisk Foundation Hallas-Møller grant to A.H. The Danish National Biobank was established with the support of major grants from the Novo Nordisk Foundation, the Danish Medical Research Council and the Lundbeck Foundation. L.S. received support from a Carlsberg Foundation postdoctoral fellowship (CF15-0899); X.L. reports funding from the Nordic Center of Excellence in Health-Related e-Sciences; K.B. received support from the Novo Nordisk Foundation (NNF14CC0001, NNF17OC0027594); D.W. reports funding from the Novo Nordisk Foundation (NNF18SA0034956, NNF14CC0001, NNF17OC0027594); T.H.P. acknowledges the Novo Nordisk Foundation (NNF18CC0034900) and the Lundbeck Foundation (R190-2014-3904); J.C. and J.W.D. report funding from the Novo Nordisk Foundation (NNF16OC0019126), The Danish Epilepsy Association, and the Central Denmark Region; C.A. is supported by The Danish National Research Foundation (Niels Bohr Professorship to John McGrath). The content is solely the responsibility of the authors and does not necessarily represent the official views of the funders.

