## Supplementary Figures and Tables for "Genome-wide association study of febrile seizures identifies seven new loci implicating fever response and neuronal excitability genes"

Skotte et al.

### Table of contents

#### Supplementary Figures


**Supplementary Figure 1:** Study design **3**

**Supplementary Figure 2:** Statistical power curves for the discovery stage analyses **4**

**Supplementary Figure 3:** Regional association plots of the seven novel febrile seizures loci **5**

**Supplementary Figure 4:** Regional association plots of all febrile seizures loci, conditional on the lead SNP at each locus **8**

**Supplementary Figure 5:** Forest plots of the seven novel febrile seizures loci **12**

**Supplementary Figure 6:** DEPICT analysis of enrichment **14**

**Supplementary Figure 7:** Forest plots of iPSYCH study subgroup results for all loci **15**

**Supplementary Figure 8:** iPSYCH study selection scheme for febrile seizures cases and controls **18**


#### Supplementary Tables


**Supplementary Table 1:** Analysis of interaction between genotype and sex at the seven novel febrile seizures loci **19**

**Supplementary Table 2:** Analysis of alternative genetic models at the seven novel febrile seizures loci **19**

**Supplementary Table 3:** Discovery stage association results for variants at the seven novel and four known febrile seizures loci **20**

**Supplementary Table 4:** eQTL results based on the BRAINEAC, DICE, eQTL Catalogue, eQTLGen, and GTEx/v8 databases for variants at all febrile seizures loci **20**

**Supplementary Table 5:** GWAS catalog associations for variants at all febrile seizures loci **20**

**Supplementary Table 6:** Febrile seizures association analysis of exome sequencing data **21**

**Supplementary Table 7:** Statistical fine mapping results for the seven novel loci **21**

**Supplementary Table 8:** Association results in the iPSYCH and DBDS cohorts for variants known to be distinctly associated with febrile seizures occurring after MMR vaccination **21**

**Supplementary Table 9:** Association results for the seven novel febrile seizures loci in the SSI discovery cohort taking MMR vaccination data into account **22**

#### Supplementary Figures

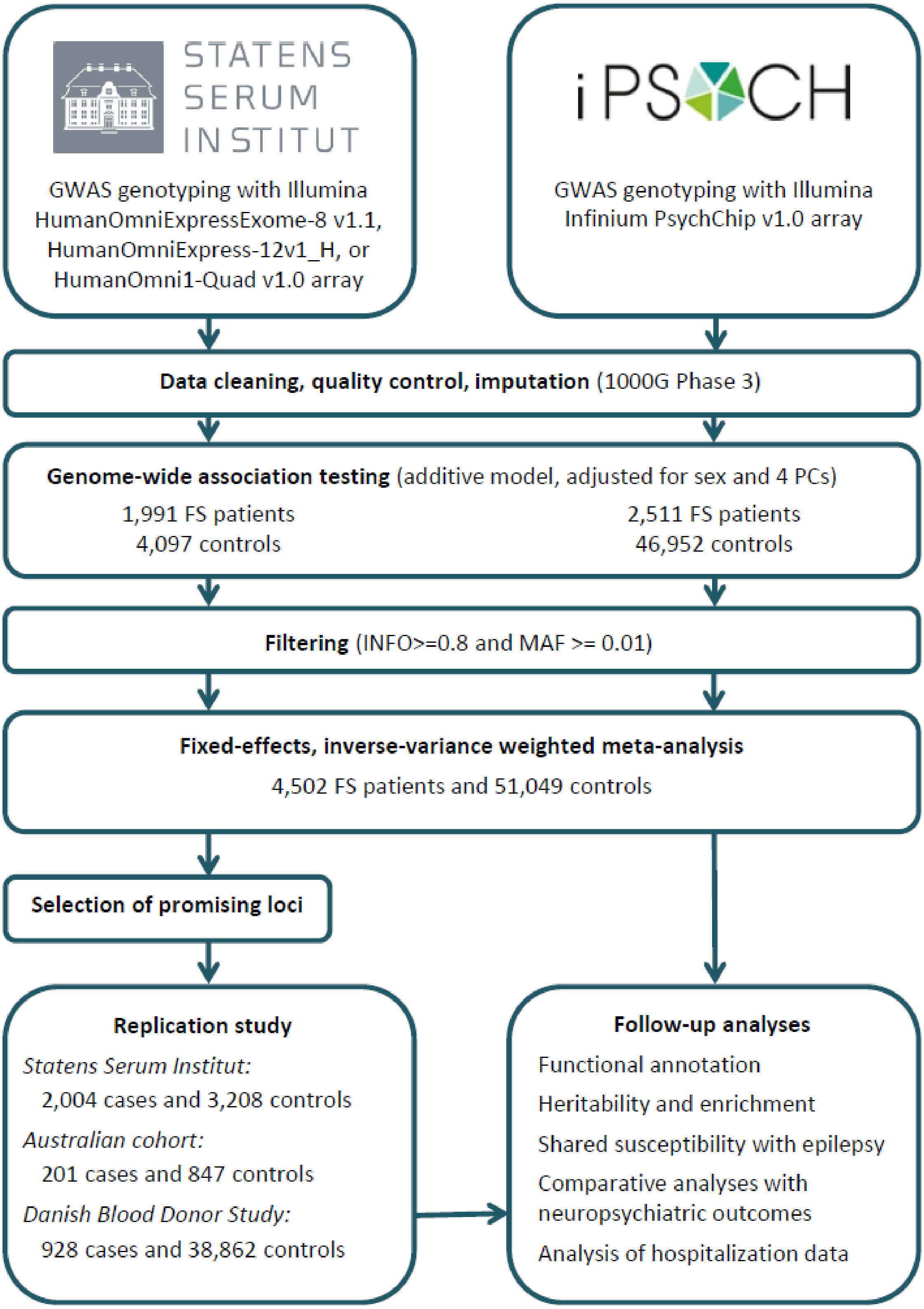


**Supplementary Figure 1.** Overview of Study Design

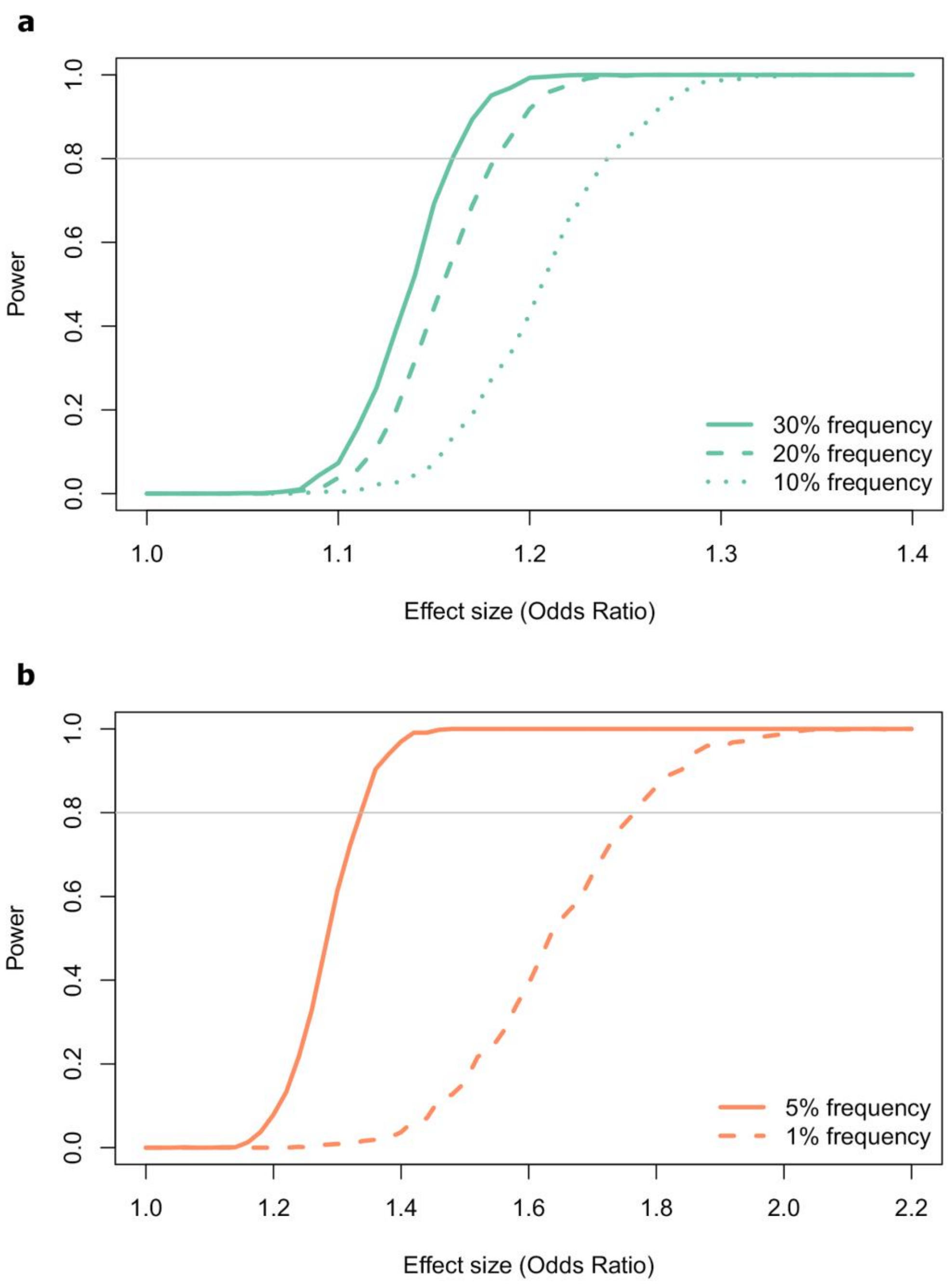


**Supplementary Figure 2.** Power simulations based on the discovery stage sample size of 4,502 cases and 51,049 controls. **a**, Simulations for variants with population risk allele frequencies of 10% (dotted line), 20% (dashed line) and 30% (full line), and **b**, for variants with frequencies of 1% (dashed line) and 5% (full line). Simulations were done assuming a febrile seizure prevalence of 3.6%, a log-additive genetic model and a significance level of $\alpha=5x10^{-8}$, for varying effect sizes. The x-axis shows the effect size in terms of the odds ratio underlying the simulation, while the y-axis shows the power (the fraction of the simulations with an α-significant *P* value for an estimated OR in the direction of the simulated OR).

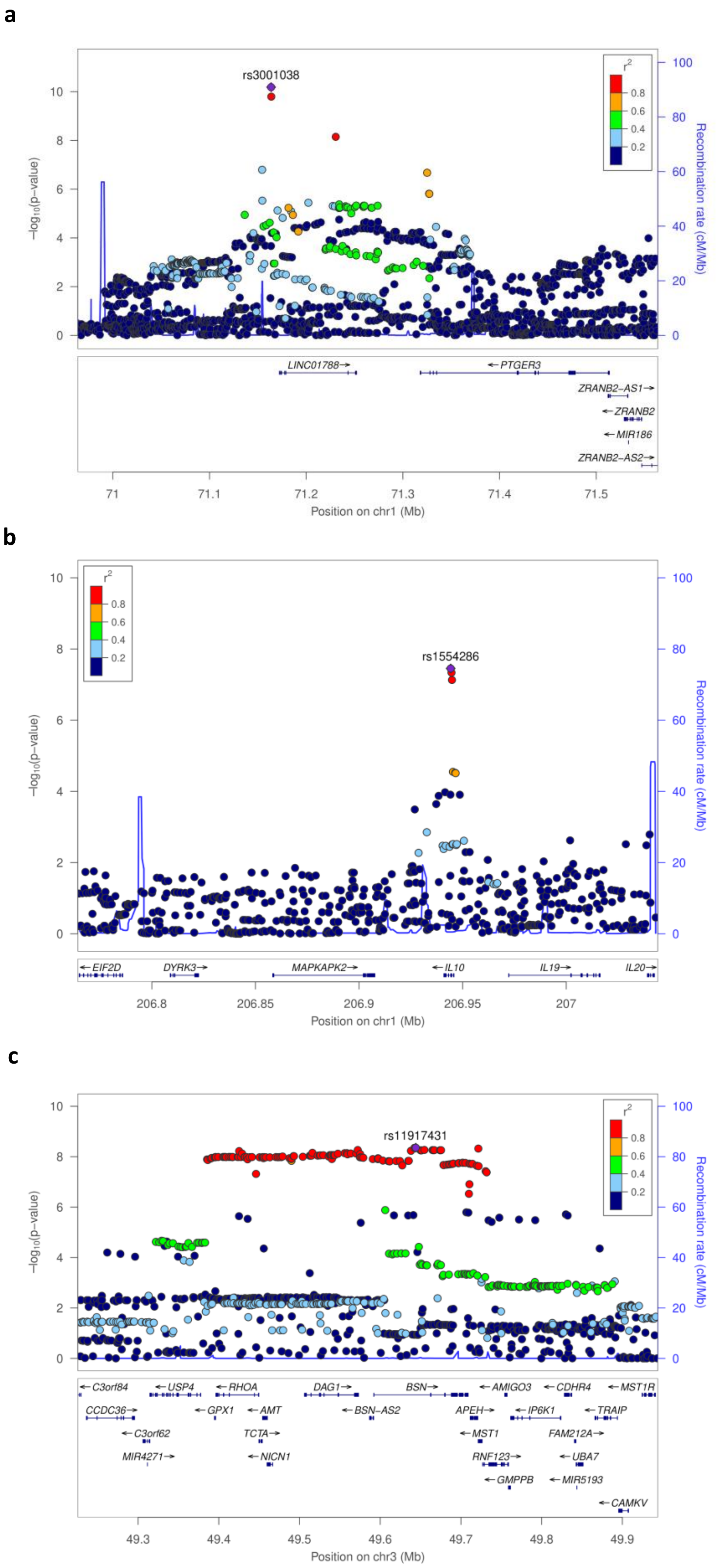
a
rs3001038
−log10(p−value)
Recombination rate (cM/Mb)
r2
LINC01788→
←PTGER3
ZRANB2−AS1→
←ZRANB2
←MIR186
ZRANB2−AS2→
Position on chr1 (Mb)
b
rs1554286
←EIF2D
DYRK3→
MAPKAPK2→
←IL10
IL19→
IL20→
Position on chr1 (Mb)
c
rs11917431
←C3orf84
←USP4
←RHOA
DAG1→
BSN→
←AMIGO3
←CDHR4
←MST1R
CCDC36→
←GPX1
←AMT
←BSN−AS2
APEH→
←IP6K1
←TRAIP
←C3orf62
TCTA→
←MST1
FAM212A→
MIR4271→
←NICN1
RNF123→
←UBA7
←GMPPB
←MIR5193
←CAMKV
Position on chr3 (Mb)

**d**

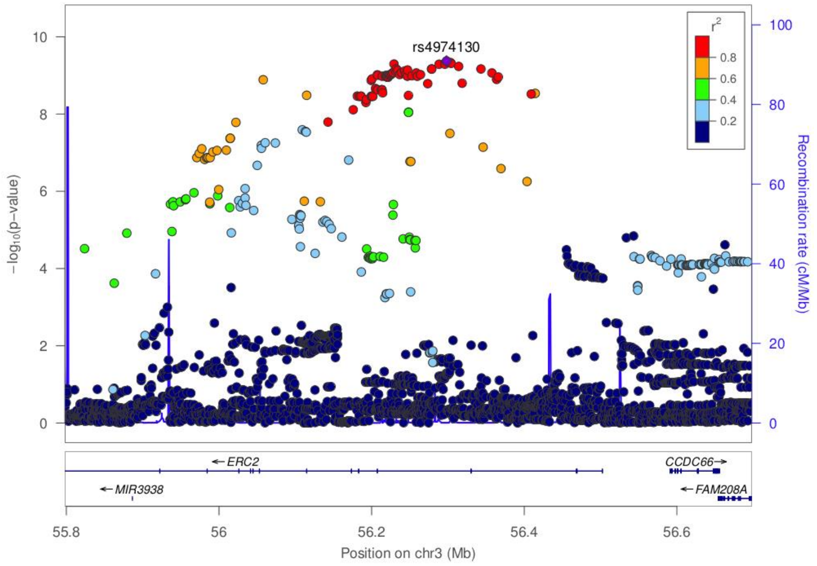
rs4974130
r2
0.8
0.6
0.4
0.2
−log10(p−value)
Recombination rate (cM/Mb)
ERC2
MIR3938
CCDC66
FAM208A
55.8
56
56.2
56.4
56.6
Position on chr3 (Mb)

**e**

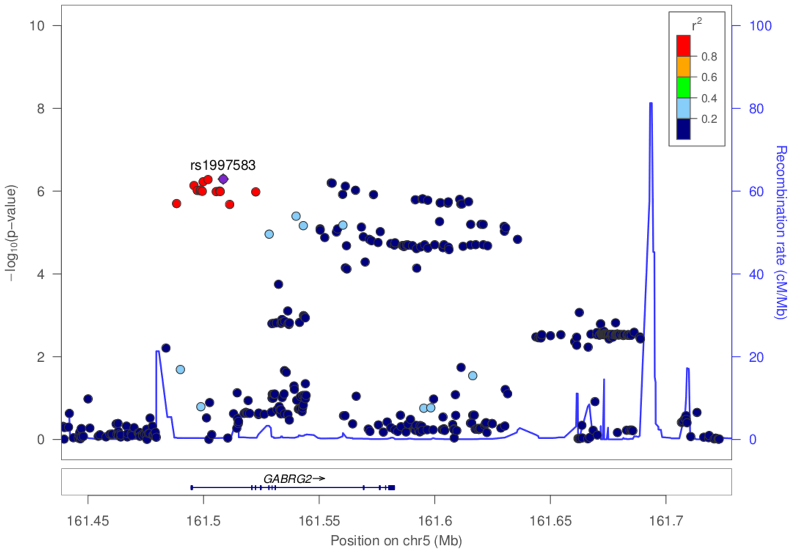
rs1997583
r2
0.8
0.6
0.4
0.2
−log10(p−value)
Recombination rate (cM/Mb)
GABRG2
161.45
161.5
161.55
161.6
161.65
161.7
Position on chr5 (Mb)

**f**

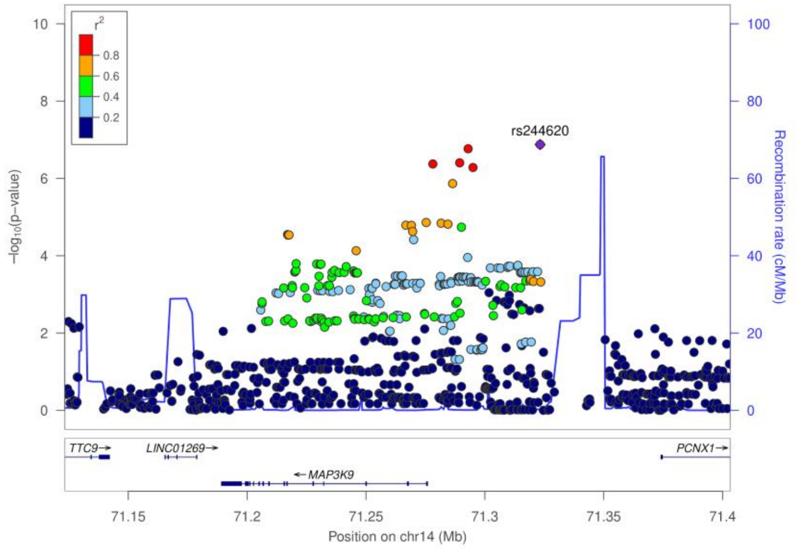
rs244620
r2
0.8
0.6
0.4
0.2
−log10(p−value)
Recombination rate (cM/Mb)
TTC9
LINC01269
MAP3K9
PCNX1
71.15
71.2
71.25
71.3
71.35
71.4
Position on chr14 (Mb)

**g**

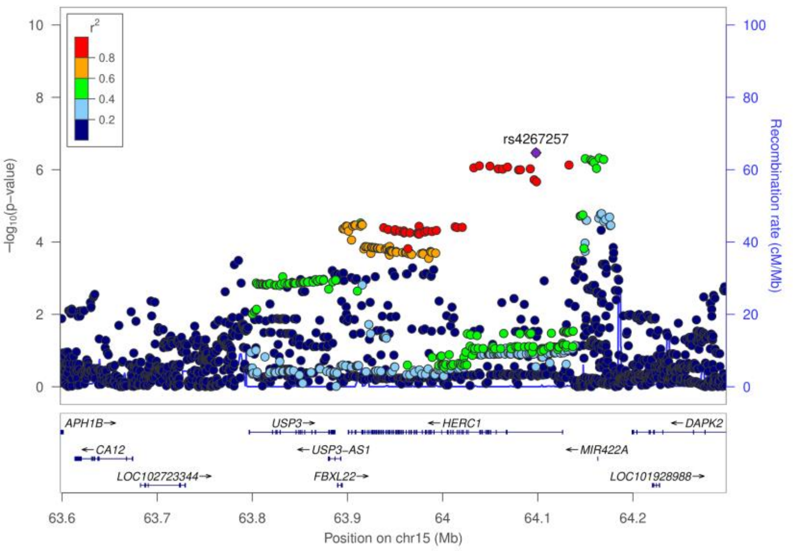


**Supplementary Figure 3.** Regional association plots of the seven novel loci for febrile seizures. Each circle represent a variant in the GWAS meta-analysis; the position along the x-axis represent the genomic location, the position along the y-axis shows the -$\log_{10}$ $P$ value, the color indicates the $r^2$ value between the variant and the lead variant at each locus. **a**, *PTGER3* locus, lead variant rs3001038, **b**, *IL10* locus, lead variant rs1554286, **c**, *BSN* locus, lead variant rs11917431, **d**, *ERC2* locus, lead variant rs4974130, **e**, *GABRG2* locus, lead variant rs1997583, **f**, *MAP3K9* locus, lead variant rs244620, and **g**, *HERC1* locus, lead variant rs4267257.

**a**

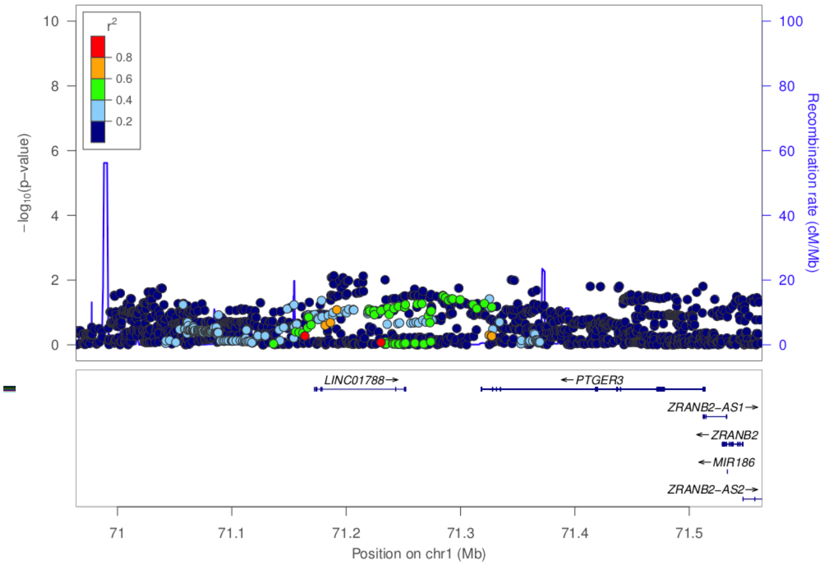
−log10(p−value)
Recombination rate (cM/Mb)
r2
0.8
0.6
0.4
0.2
LINC01788→
←PTGER3
ZRANB2−AS1→
←ZRANB2
←MIR186
ZRANB2−AS2→
71
71.1
71.2
71.3
71.4
71.5
Position on chr1 (Mb)

**b**

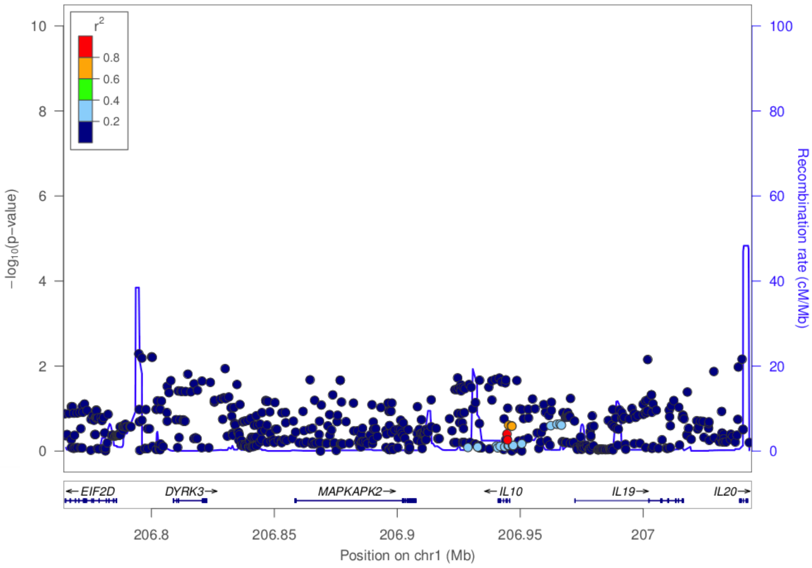
−log10(p−value)
Recombination rate (cM/Mb)
r2
0.8
0.6
0.4
0.2
←EIF2D
DYRK3→
MAPKAPK2→
←IL10
IL19→
IL20→
206.8
206.85
206.9
206.95
207
Position on chr1 (Mb)

**c**

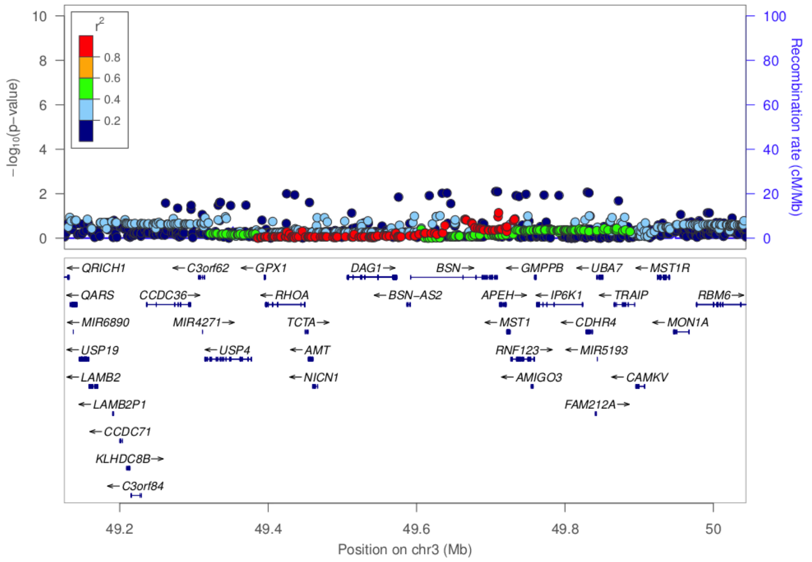
−log10(p−value)
Recombination rate (cM/Mb)
r2
0.8
0.6
0.4
0.2
←QRICH1
←QARS
←MIR6890
←USP19
←LAMB2
←LAMB2P1
←CCDC71
KLHDC8B→
←C3orf84
←C3orf62
CCDC36→
MIR4271→
←USP4
←GPX1
←RHOA
TCTA→
←AMT
←NICN1
DAG1→
BSN→
←BSN−AS2
←GMPPB
APEH→
←MST1
RNF123→
←AMIGO3
←UBA7
←IP6K1
←CDHR4
←MIR5193
FAM212A→
←MST1R
←TRAIP
←MON1A
←CAMKV
RBM6→
49.2
49.4
49.6
49.8
50
Position on chr3 (Mb)

**d**

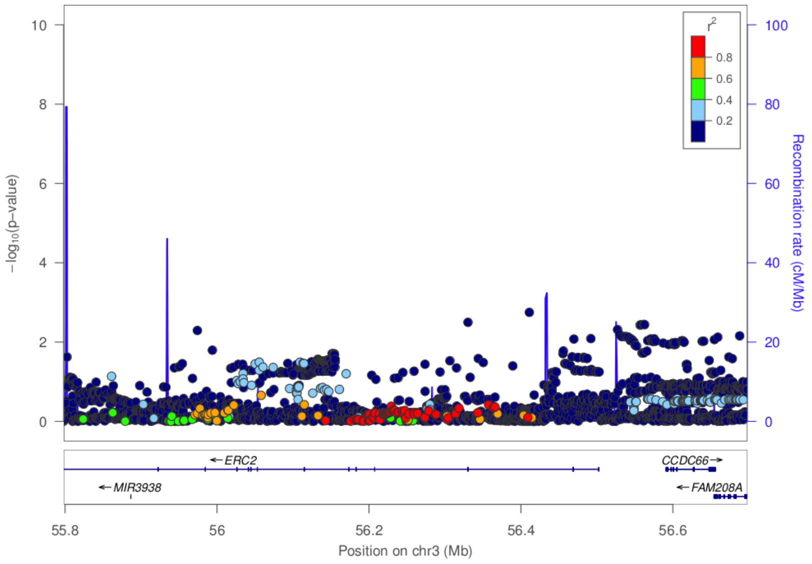
−log10(p−value)
Recombination rate (cM/Mb)
r2
0.8
0.6
0.4
0.2
←ERC2
CCDC66→
←MIR3938
←FAM208A
55.8
56
56.2
56.4
56.6
Position on chr3 (Mb)

**e**

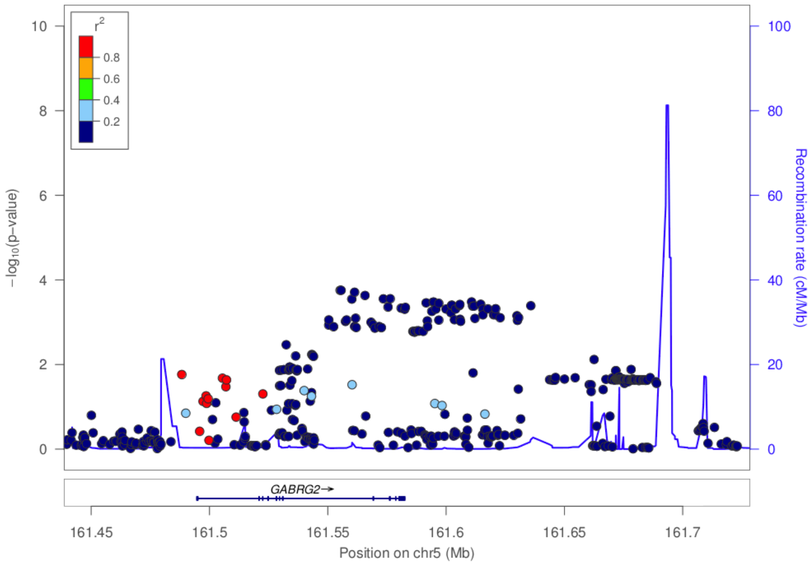
−log10(p−value)
Recombination rate (cM/Mb)
r2
0.8
0.6
0.4
0.2
GABRG2→
161.45
161.5
161.55
161.6
161.65
161.7
Position on chr5 (Mb)

**f**

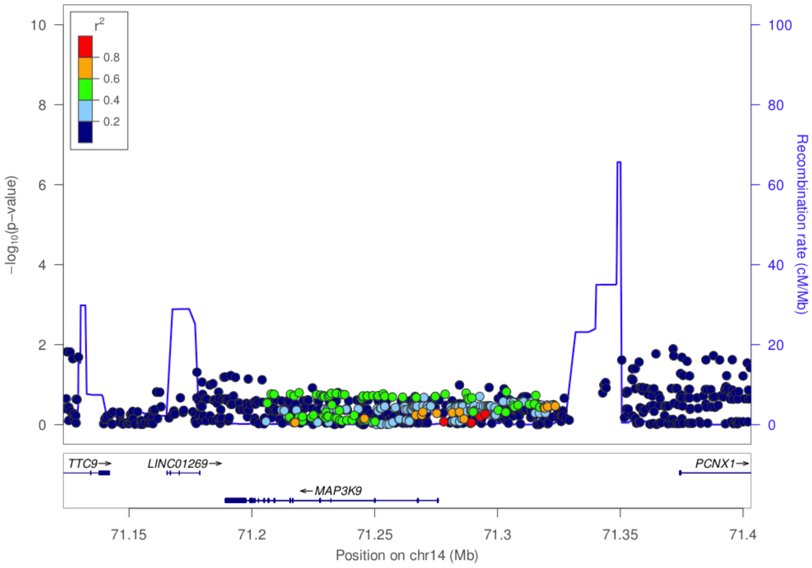
−log10(p−value)
Recombination rate (cM/Mb)
r2
0.8
0.6
0.4
0.2
TTC9→
LINC01269→
←MAP3K9
PCNX1→
71.15
71.2
71.25
71.3
71.35
71.4
Position on chr14 (Mb)

**g**

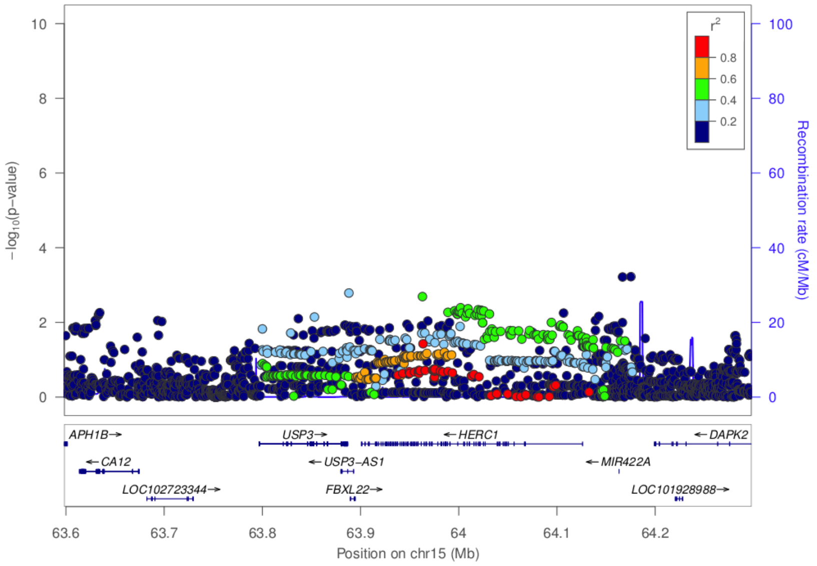

r²
0.8
0.6
0.4
0.2
−log10(p−value)
Recombination rate (cM/Mb)
APH1B→
CA12
LOC102723344→
USP3→
USP3−AS1
FBXL22→
HERC1
MIR422A
DAPK2
LOC101928988→
Position on chr15 (Mb)


**h**

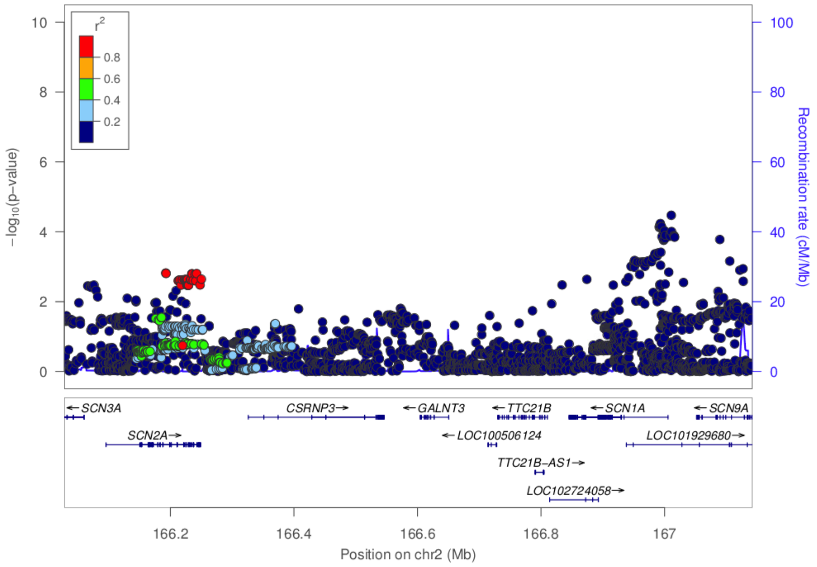

r²
0.8
0.6
0.4
0.2
−log10(p−value)
Recombination rate (cM/Mb)
SCN3A
SCN2A→
CSRNP3→
GALNT3
LOC100506124
TTC21B
TTC21B−AS1→
LOC102724058→
SCN1A
SCN9A
LOC101929680→
Position on chr2 (Mb)


**i**

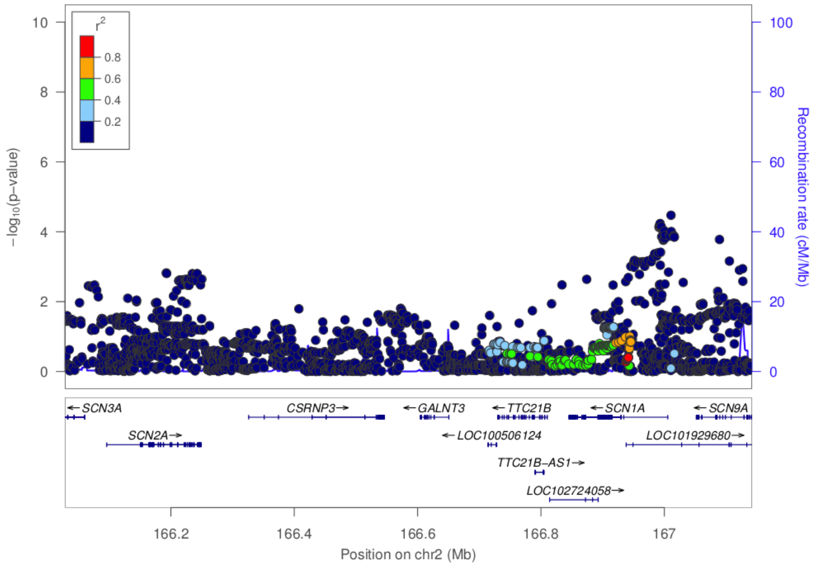

r²
0.8
0.6
0.4
0.2
−log10(p−value)
Recombination rate (cM/Mb)
SCN3A
SCN2A→
CSRNP3→
GALNT3
LOC100506124
TTC21B
TTC21B−AS1→
LOC102724058→
SCN1A
SCN9A
LOC101929680→
Position on chr2 (Mb)

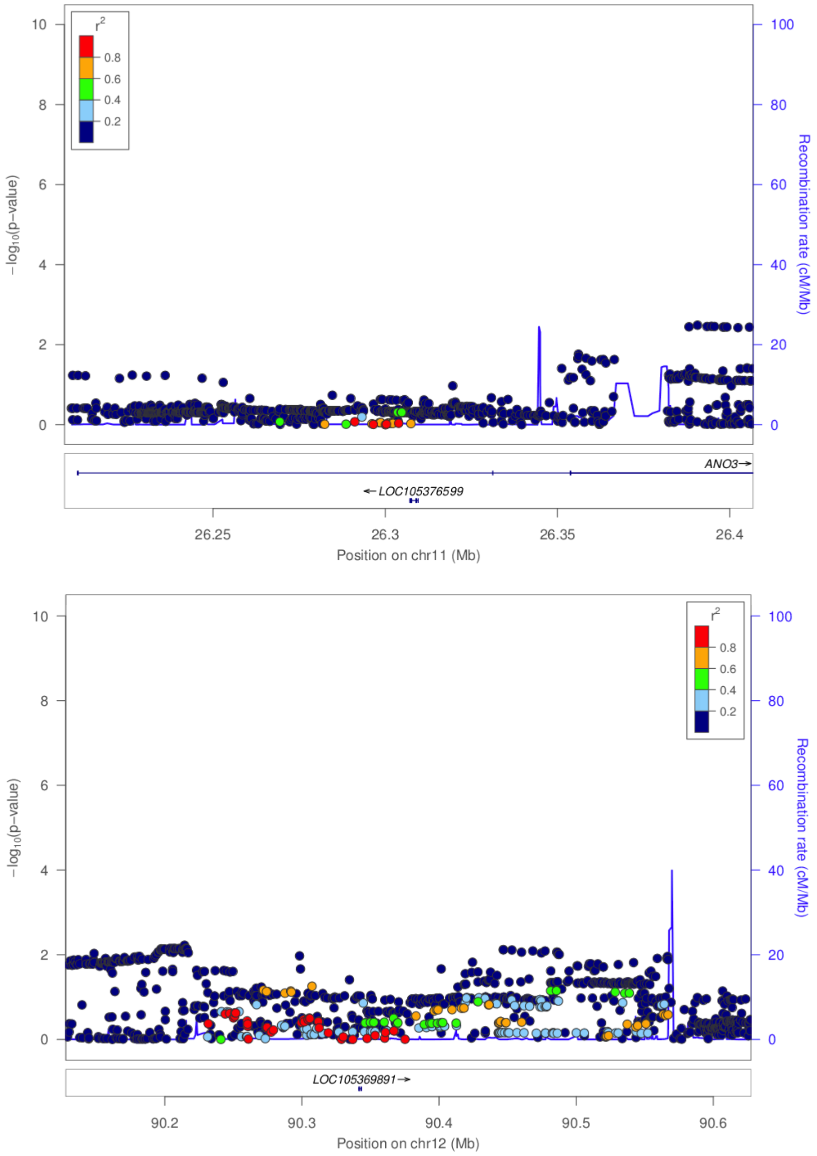


**Supplementary Figure 4.** Regional association plots of the seven novel and four known loci for febrile seizures, conditioning on the lead SNP at each locus. Each circle represent a variant in the GWAS meta-analysis; the position along the x-axis represent the genomic location, the position along the y-axis shows the -$\log_{10}$ *P* value, the color indicates the $r^2$ value between the variant and the lead variant at each locus. **a**, *PTGER3* locus, conditioned on lead variant rs3001038, **b**, *IL10* locus, conditioned on lead variant rs1554286, **c**, *BSN* locus, conditioned on lead variant rs11917431, **d**, *ERC2* locus, conditioned on lead variant rs4974130, **e**, *GABRG2* locus, conditioned on lead variant rs1997583, **f**, *MAP3K9* locus, conditioned on lead variant rs244620, **g**, *HERC1* locus, conditioned on lead variant rs4267257, **h**, *SCN2A* locus, conditioned on lead variant rs16850528 and neighboring *SCN1A* lead variant rs11904006, **i**, *SCN1A* locus, conditioned on lead variant rs11904006 and neighboring *SCN2A* lead variant rs16850528, **j**, *ANO3* locus, conditioned on lead variant rs114444506, **k**, 12q21.33 locus, conditioned on lead variant rs71909799.

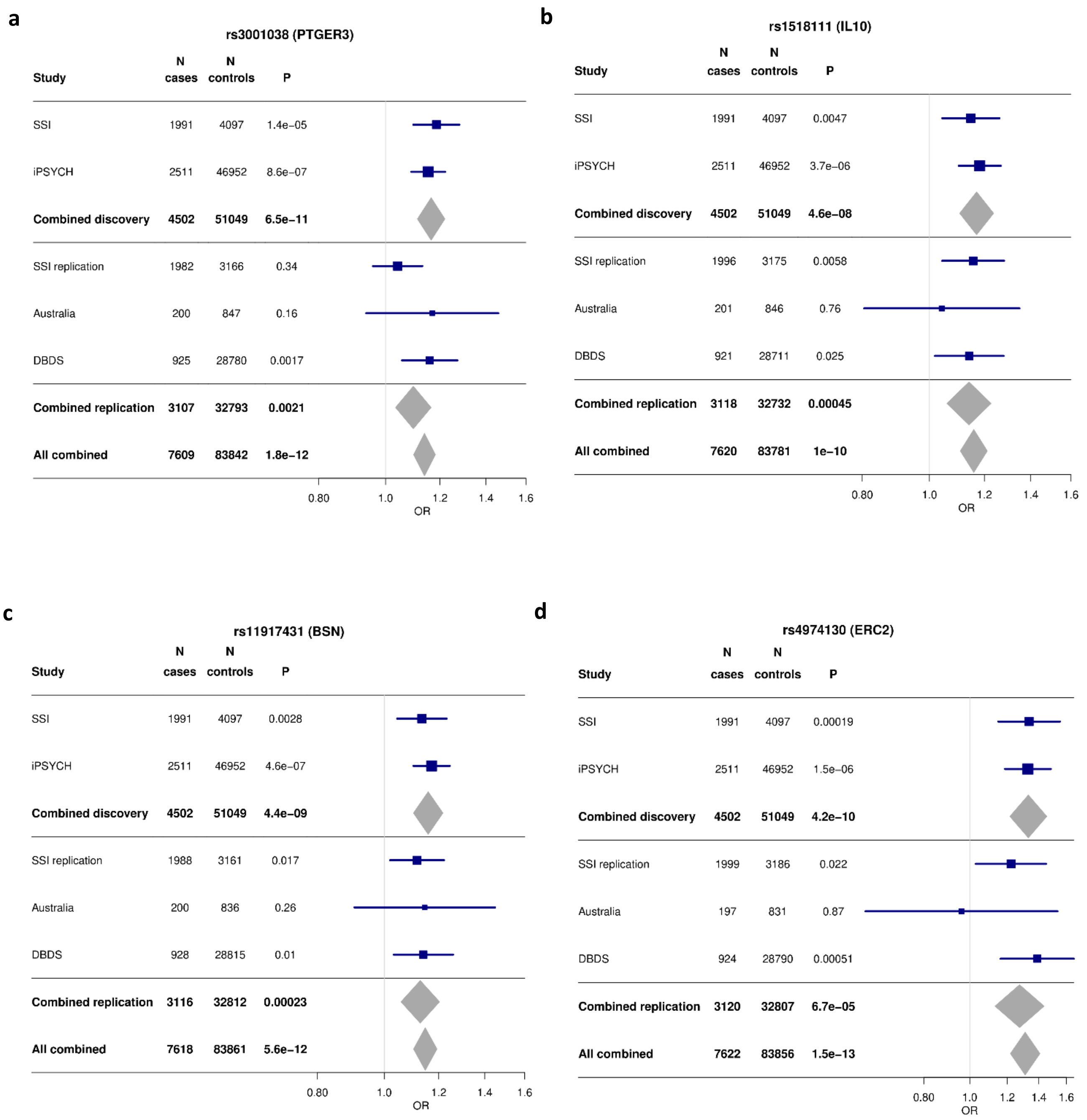

a
rs3001038 (PTGER3)
Study | N cases | N controls | P
SSI 1991 4097 1.4e−05
iPSYCH 2511 46952 8.6e−07
Combined discovery 4502 51049 6.5e−11
SSI replication 1982 3166 0.34
Australia 200 847 0.16
DBDS 925 28780 0.0017
Combined replication 3107 32793 0.0021
All combined 7609 83842 1.8e−12
0.80 1.0 1.2 1.4 1.6
OR
b
rs1518111 (IL10)
Study | N cases | N controls | P
SSI 1991 4097 0.0047
iPSYCH 2511 46952 3.7e−06
Combined discovery 4502 51049 4.6e−08
SSI replication 1996 3175 0.0058
Australia 201 846 0.76
DBDS 921 28711 0.025
Combined replication 3118 32732 0.00045
All combined 7620 83781 1e−10
0.80 1.0 1.2 1.4 1.6
OR
c
rs11917431 (BSN)
Study | N cases | N controls | P
SSI 1991 4097 0.0028
iPSYCH 2511 46952 4.6e−07
Combined discovery 4502 51049 4.4e−09
SSI replication 1988 3161 0.017
Australia 200 836 0.26
DBDS 928 28815 0.01
Combined replication 3116 32812 0.00023
All combined 7618 83861 5.6e−12
0.80 1.0 1.2 1.4 1.6
OR
d
rs4974130 (ERC2)
Study | N cases | N controls | P
SSI 1991 4097 0.00019
iPSYCH 2511 46952 1.5e−06
Combined discovery 4502 51049 4.2e−10
SSI replication 1999 3186 0.022
Australia 197 831 0.87
DBDS 924 28790 0.00051
Combined replication 3120 32807 6.7e−05
All combined 7622 83856 1.5e−13
0.80 1.0 1.2 1.4 1.6
OR

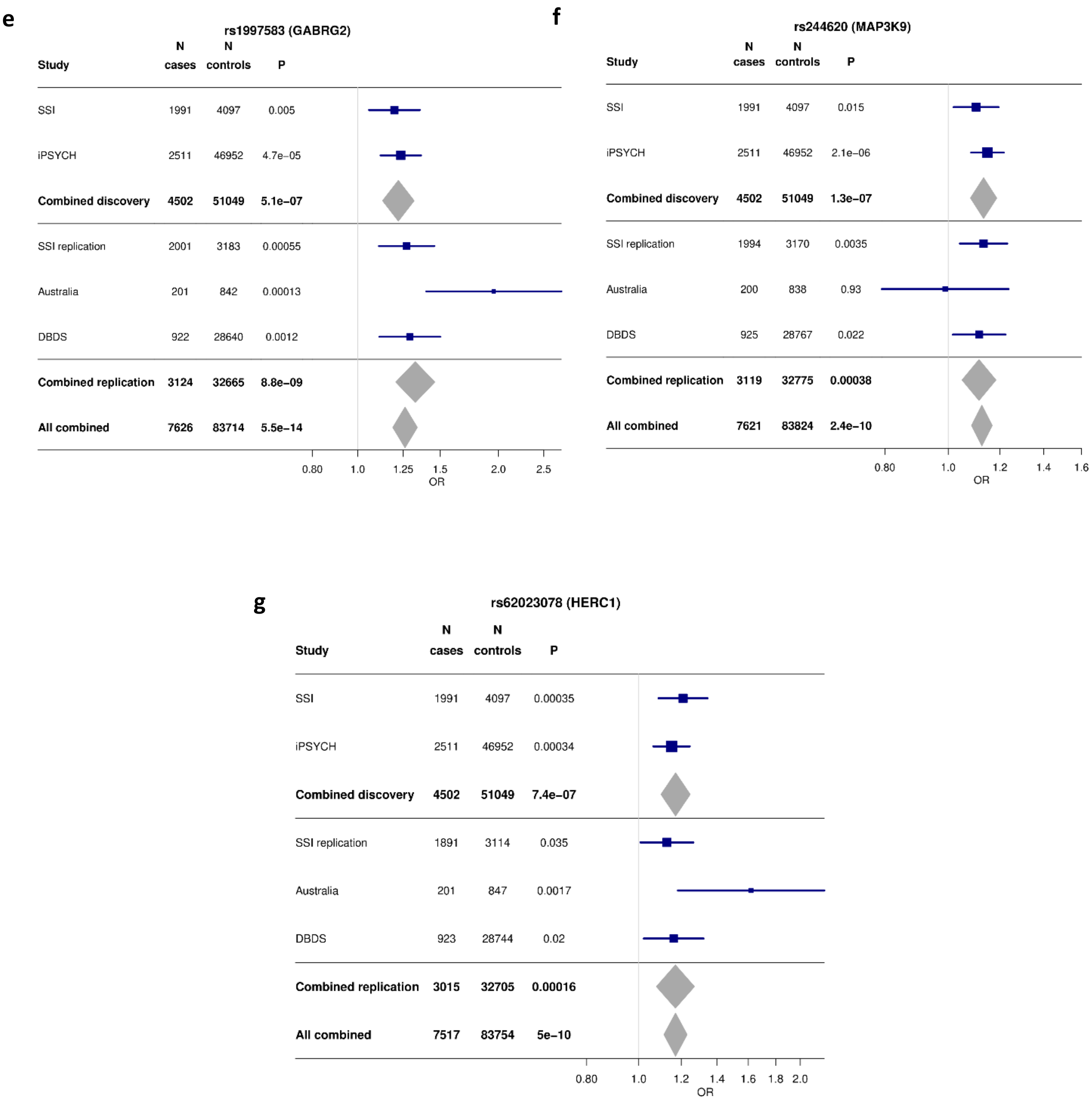


**Supplementary Figure 5.** Forest plots showing associations between the robustly replicating SNP at each of the seven novel loci and febrile seizures in contributing cohorts and in combined analyses. The plots show odds ratio estimates with 95% confidence intervals. **a**, *PTGER3* locus, rs3001038, **b**, *IL10* locus, rs1518111, **c**, *BSN* locus, rs11917431, **d**, *ERC2* locus, rs4974130, **e**, *GABRG2* locus, rs1997583, **f**, *MAP3K9* locus, rs244620, and **g**, *HERC1* locus, rs62023078.

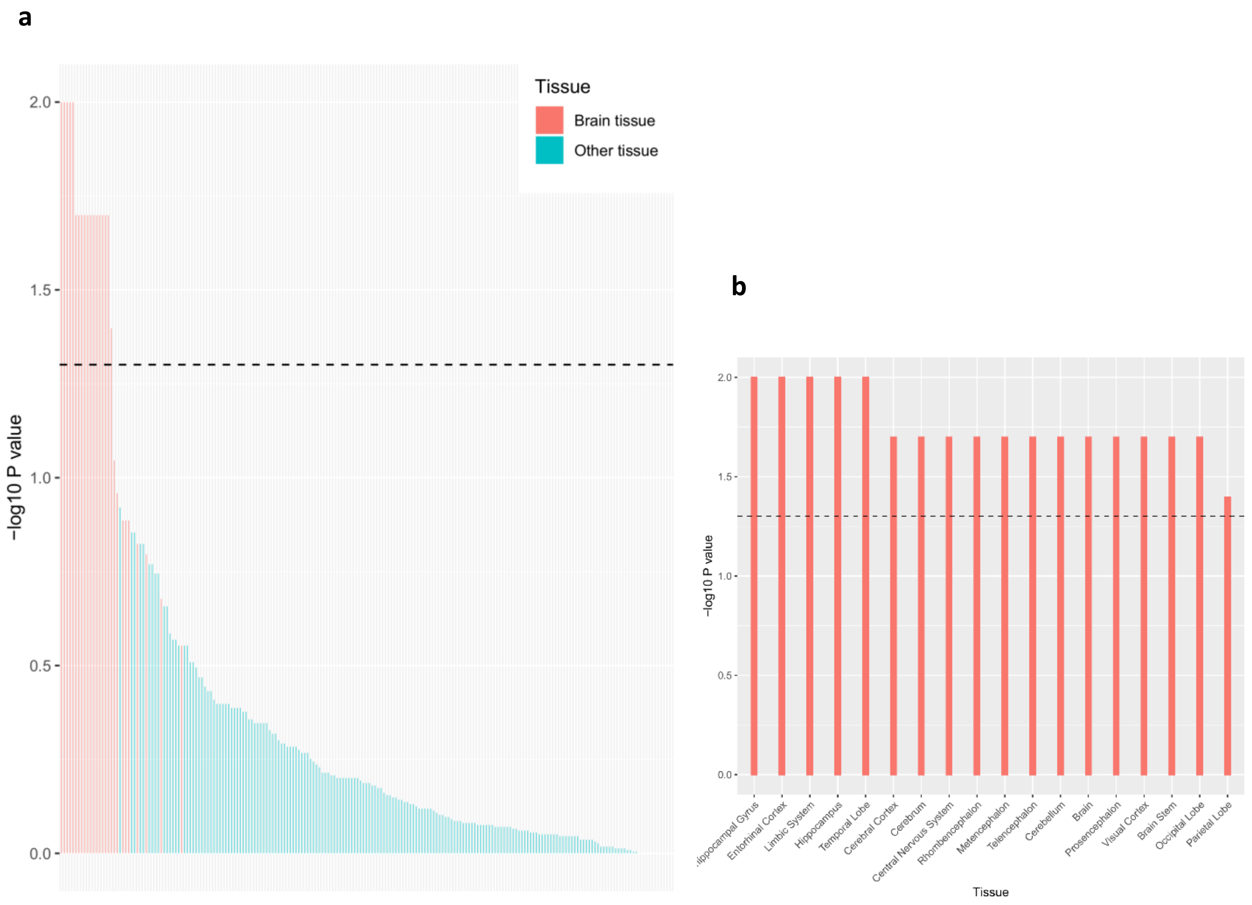


**Supplementary Figure 6.** DEPICT analysis of enrichment. **a**, overall results for 209 tissue and cell types with brain tissues highlighted. The dashed line corresponds to $P = 0.05$, and **b** shows results for tissues with $P < 0.05$.

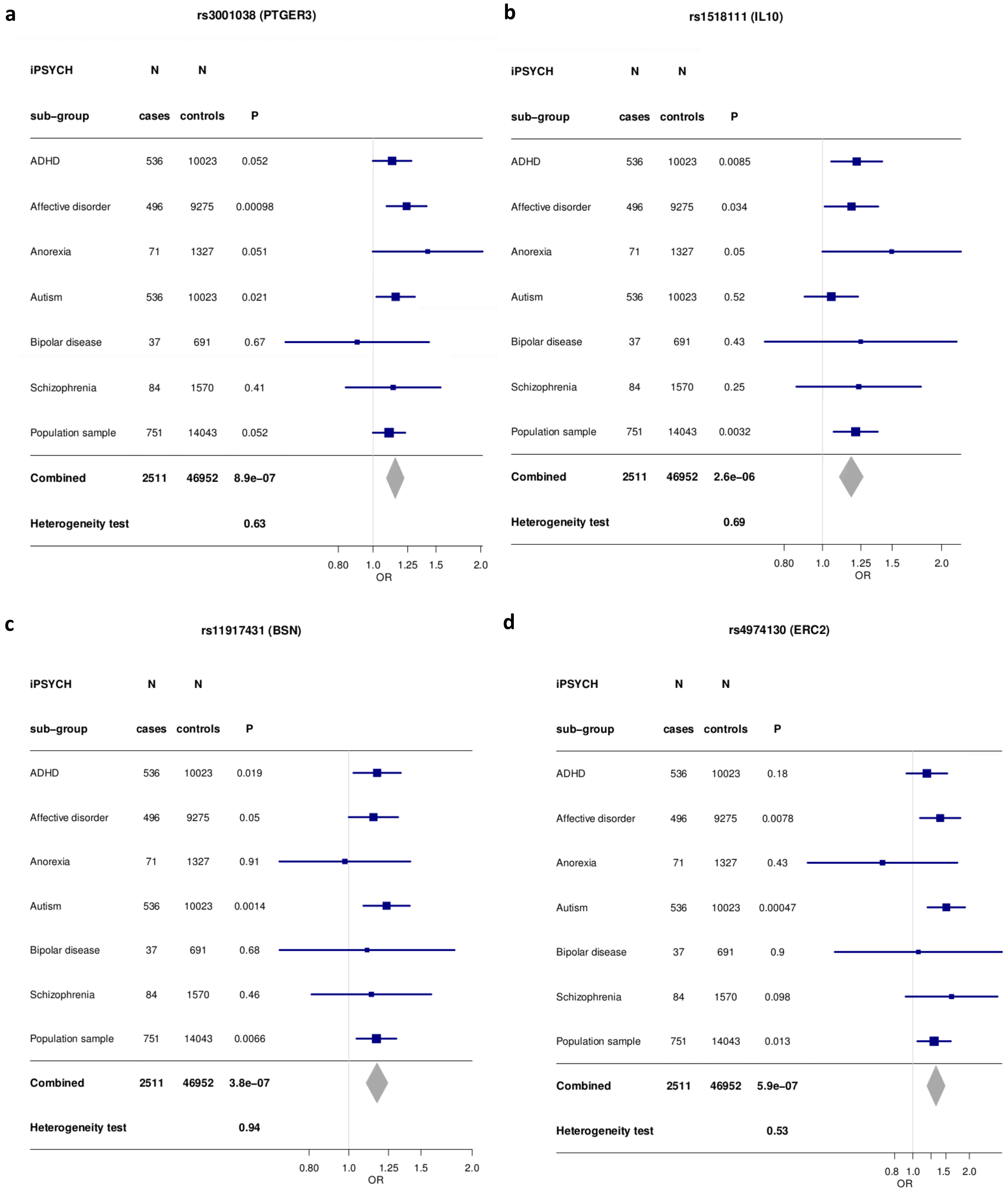
a
rs3001038 (PTGER3)
iPSYCH sub–group | N cases | N controls | P
ADHD 536 10023 0.052
Affective disorder 496 9275 0.00098
Anorexia 71 1327 0.051
Autism 536 10023 0.021
Bipolar disease 37 691 0.67
Schizophrenia 84 1570 0.41
Population sample 751 14043 0.052
Combined 2511 46952 8.9e–07
Heterogeneity test 0.63
0.80 1.0 1.25 1.5 2.0
OR
b
rs1518111 (IL10)
iPSYCH sub–group | N cases | N controls | P
ADHD 536 10023 0.0085
Affective disorder 496 9275 0.034
Anorexia 71 1327 0.05
Autism 536 10023 0.52
Bipolar disease 37 691 0.43
Schizophrenia 84 1570 0.25
Population sample 751 14043 0.0032
Combined 2511 46952 2.6e–06
Heterogeneity test 0.69
0.80 1.0 1.25 1.5 2.0
OR
c
rs11917431 (BSN)
iPSYCH sub–group | N cases | N controls | P
ADHD 536 10023 0.019
Affective disorder 496 9275 0.05
Anorexia 71 1327 0.91
Autism 536 10023 0.0014
Bipolar disease 37 691 0.68
Schizophrenia 84 1570 0.46
Population sample 751 14043 0.0066
Combined 2511 46952 3.8e–07
Heterogeneity test 0.94
0.80 1.0 1.25 1.5 2.0
OR
d
rs4974130 (ERC2)
iPSYCH sub–group | N cases | N controls | P
ADHD 536 10023 0.18
Affective disorder 496 9275 0.0078
Anorexia 71 1327 0.43
Autism 536 10023 0.00047
Bipolar disease 37 691 0.9
Schizophrenia 84 1570 0.098
Population sample 751 14043 0.013
Combined 2511 46952 5.9e–07
Heterogeneity test 0.53
0.8 1.0 1.5 2.0
OR

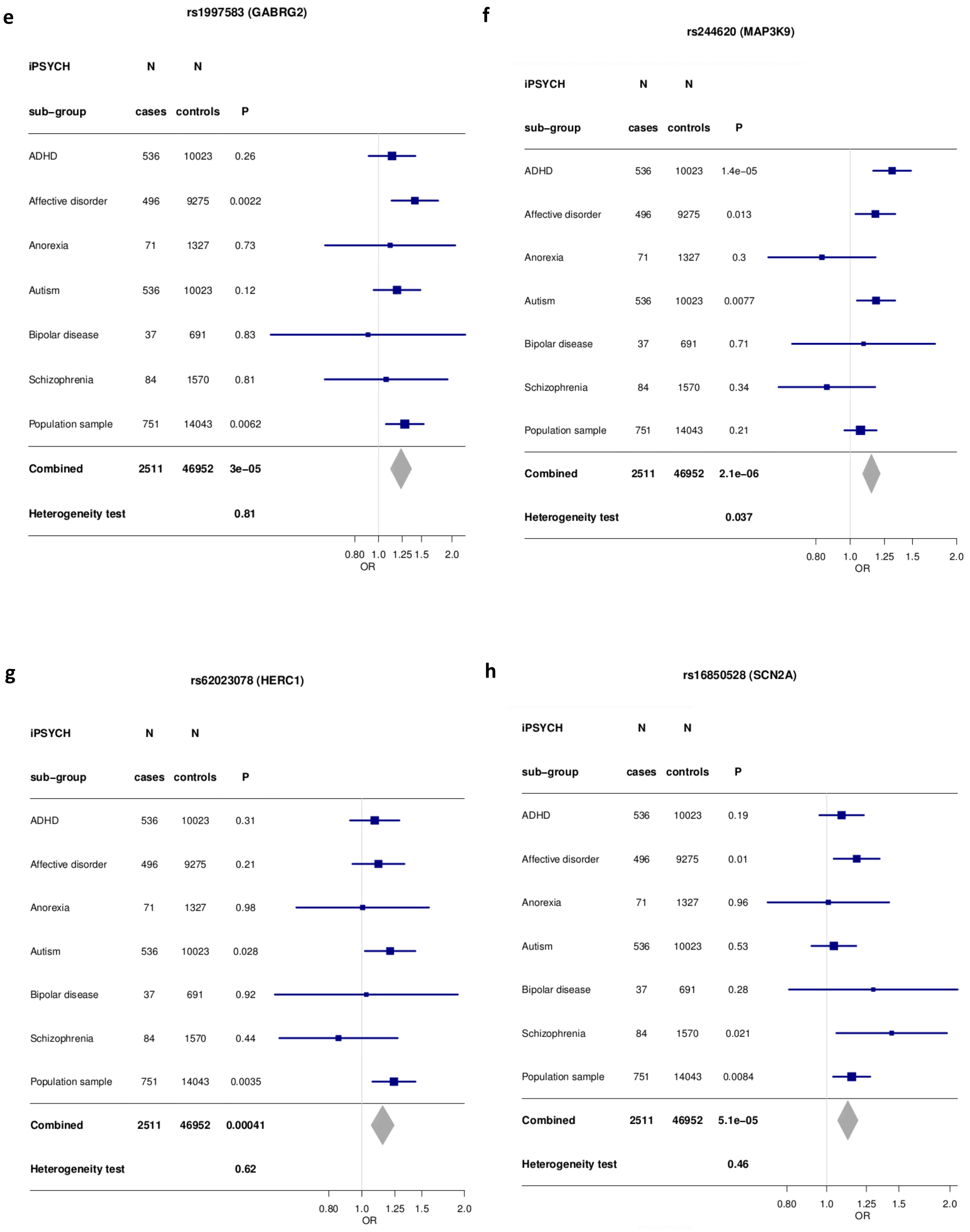
e
rs1997583 (GABRG2)
iPSYCH sub–group | N cases | N controls | P
ADHD 536 10023 0.26
Affective disorder 496 9275 0.0022
Anorexia 71 1327 0.73
Autism 536 10023 0.12
Bipolar disease 37 691 0.83
Schizophrenia 84 1570 0.81
Population sample 751 14043 0.0062
Combined 2511 46952 3e–05
Heterogeneity test 0.81
0.80 1.0 1.25 1.5 2.0
OR
f
rs244620 (MAP3K9)
iPSYCH sub–group | N cases | N controls | P
ADHD 536 10023 1.4e–05
Affective disorder 496 9275 0.013
Anorexia 71 1327 0.3
Autism 536 10023 0.0077
Bipolar disease 37 691 0.71
Schizophrenia 84 1570 0.34
Population sample 751 14043 0.21
Combined 2511 46952 2.1e–06
Heterogeneity test 0.037
0.80 1.0 1.25 1.5 2.0
OR
g
rs62023078 (HERC1)
iPSYCH sub–group | N cases | N controls | P
ADHD 536 10023 0.31
Affective disorder 496 9275 0.21
Anorexia 71 1327 0.98
Autism 536 10023 0.028
Bipolar disease 37 691 0.92
Schizophrenia 84 1570 0.44
Population sample 751 14043 0.0035
Combined 2511 46952 0.00041
Heterogeneity test 0.62
0.80 1.0 1.25 1.5 2.0
OR
h
rs16850528 (SCN2A)
iPSYCH sub–group | N cases | N controls | P
ADHD 536 10023 0.19
Affective disorder 496 9275 0.01
Anorexia 71 1327 0.96
Autism 536 10023 0.53
Bipolar disease 37 691 0.28
Schizophrenia 84 1570 0.021
Population sample 751 14043 0.0084
Combined 2511 46952 5.1e–05
Heterogeneity test 0.46
0.80 1.0 1.25 1.5 2.0
OR

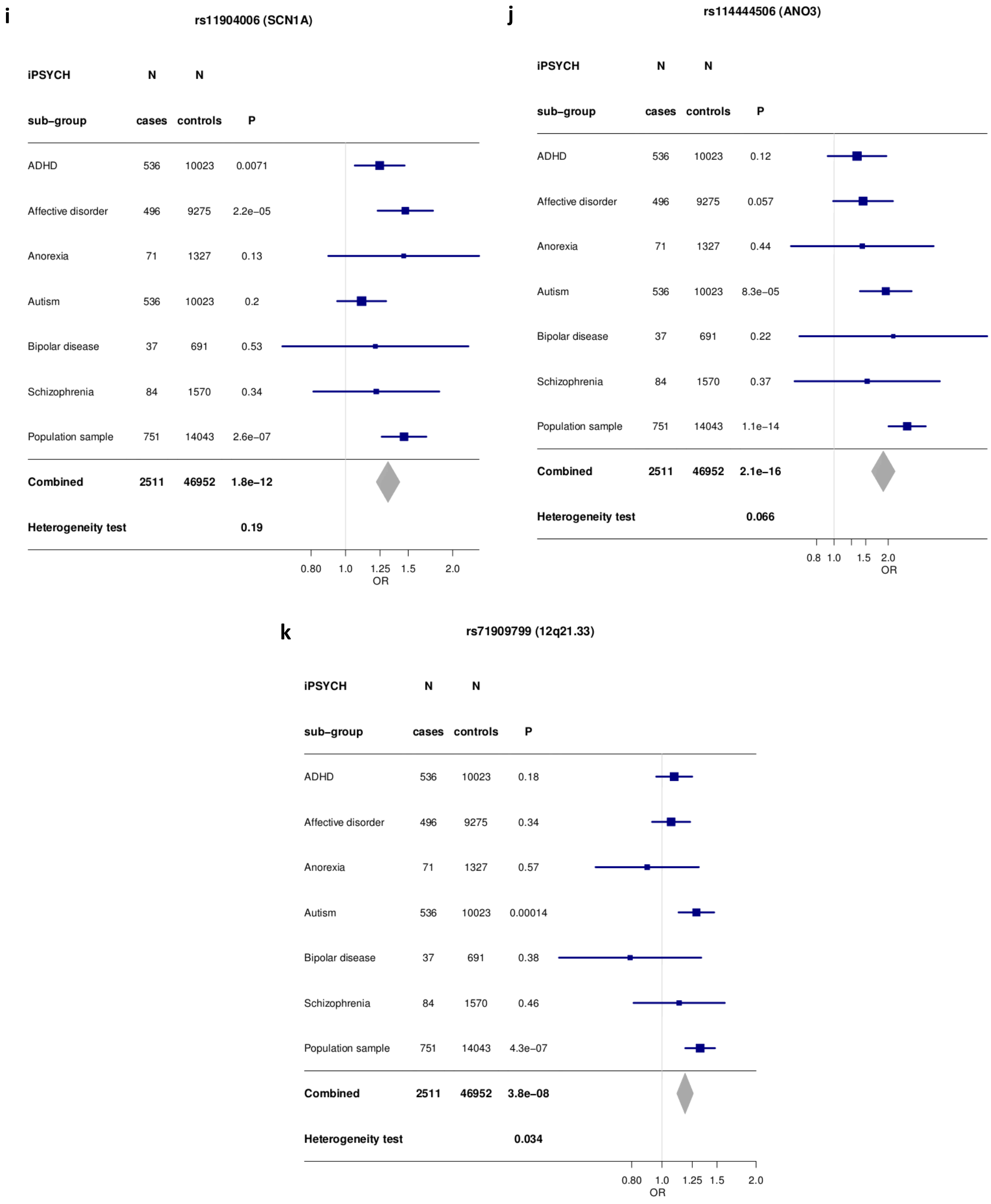


**Supplementary Figure 7.** Analyses of heterogeneity between subgroups of the iPSYCH study for the seven novel and four known febrile seizures loci. The forest plots show odds ratio estimates with 95% confidence intervals. **a**, *PTGER3* locus, rs3001038, **b**, *IL10* locus, rs1554286, **c**, *BSN* locus, rs11917431, **d**, *ERC2* locus, rs4974130, **e**, *GABRG2* locus, rs1997583, **f**, *MAP3K9* locus, rs244620, **g**, *HERC1* locus, rs4267257, **h**, *SCN2A* locus, rs16850528, **i**, *SCN1A* locus, rs11904006, **j**, *ANO3* locus, rs114444506, **k**, 12q21.33 locus, rs71909799.

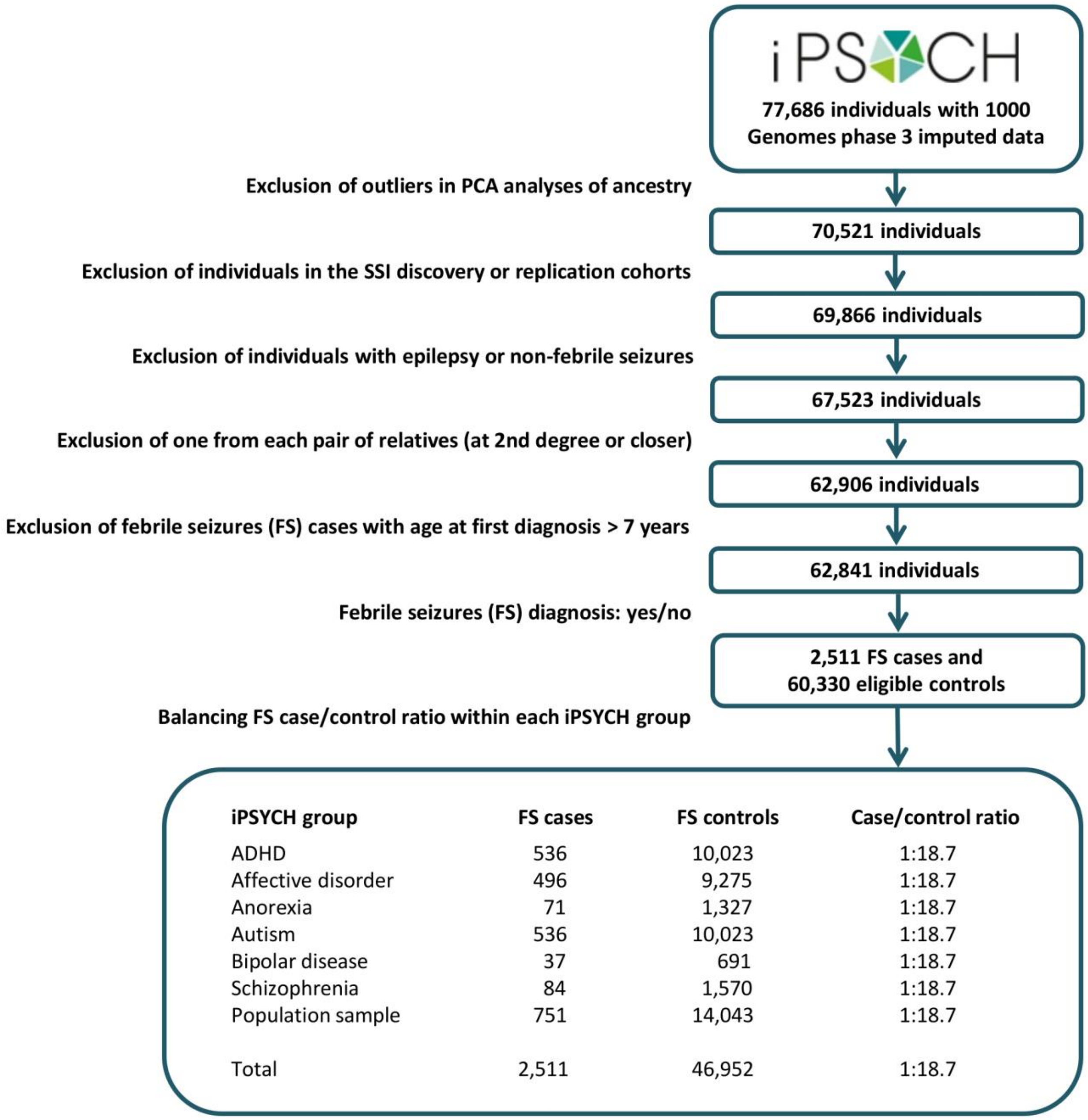


| iPSYCH group | FS cases | FS controls | Case/control ratio |
|---|---|---|---|
| ADHD | 536 | 10,023 | 1:18.7 |
| Affective disorder | 496 | 9,275 | 1:18.7 |
| Anorexia | 71 | 1,327 | 1:18.7 |
| Autism | 536 | 10,023 | 1:18.7 |
| Bipolar disease | 37 | 691 | 1:18.7 |
| Schizophrenia | 84 | 1,570 | 1:18.7 |
| Population sample | 751 | 14,043 | 1:18.7 |
| Total | 2,511 | 46,952 | 1:18.7 |

**Supplementary Figure 8.** Selection of febrile seizures cases and controls in the iPSYCH study.

### Supplementary Tables

#### Supplementary Table 1

Analysis of interaction between genotype and sex for the seven novel febrile seizures loci in the SSI and iPSYCH discovery cohorts individually and combined by meta-analysis.

| Chr | Position | SNP | Allele1 | Allele2 | Sample_set | Beta_SNP | SE_SNP | P_SNP | Beta_sex | SE_sex | P_sex | Beta_int | SE_int | P_int |
|---|---|---|---|---|---|---|---|---|---|---|---|---|---|---|
| 1 | 71163715 | rs3001038 | A | G | SSI | 0.097 | 0.054 | 0.0712 | -0.247 | 0.098 | 0.012 | 0.154 | 0.079 | 0.05 |
| | | | | | iPSYCH | 0.132 | 0.038 | 0.000442 | -0.179 | 0.075 | 0.017 | 0.027 | 0.059 | 0.648 |
| | | | | | Meta | | | | | | | 0.073 | 0.047 | 0.122 |
| 1 | 206944645 | rs1518111 | T | C | SSI | 0.141 | 0.067 | 0.0356 | -0.093 | 0.068 | 0.172 | -0.007 | 0.097 | 0.942 |
| | | | | | iPSYCH | 0.177 | 0.046 | 0.000112 | -0.143 | 0.052 | 0.006 | -0.025 | 0.073 | 0.733 |
| | | | | | Meta | | | | | | | -0.018 | 0.058 | 0.751 |
| 3 | 49644012 | rs11917431 | T | C | SSI | 0.061 | 0.058 | 0.293 | -0.176 | 0.075 | 0.019 | 0.142 | 0.085 | 0.094 |
| | | | | | iPSYCH | 0.147 | 0.04 | 0.000281 | -0.173 | 0.058 | 0.003 | 0.03 | 0.064 | 0.638 |
| | | | | | Meta | | | | | | | 0.07 | 0.051 | 0.167 |
| 3 | 56298138 | rs4974130 | C | T | SSI | 0.361 | 0.105 | 0.000556 | -0.073 | 0.059 | 0.212 | -0.155 | 0.154 | 0.314 |
| | | | | | iPSYCH | 0.314 | 0.073 | 1.71E-05 | -0.143 | 0.045 | 0.001 | -0.078 | 0.116 | 0.505 |
| | | | | | Meta | | | | | | | -0.106 | 0.093 | 0.255 |
| 5 | 161508503 | rs1997583 | T | A | SSI | 0.239 | 0.09 | 0.00763 | -0.072 | 0.061 | 0.24 | -0.119 | 0.128 | 0.353 |
| | | | | | iPSYCH | 0.223 | 0.066 | 0.000675 | -0.148 | 0.047 | 0.002 | -0.028 | 0.104 | 0.79 |
| | | | | | Meta | | | | | | | -0.064 | 0.081 | 0.428 |
| 14 | 71323249 | rs244620 | C | T | SSI | 0.053 | 0.056 | 0.343 | -0.181 | 0.093 | 0.051 | 0.096 | 0.081 | 0.235 |
| | | | | | iPSYCH | 0.158 | 0.038 | 2.66E-05 | -0.105 | 0.071 | 0.138 | -0.05 | 0.059 | 0.404 |
| | | | | | Meta | | | | | | | 0.001 | 0.048 | 0.978 |
| 15 | 64132818 | rs62023078 | G | T | SSI | 0.237 | 0.074 | 0.00133 | 0.074 | 0.189 | 0.693 | -0.101 | 0.107 | 0.349 |
| | | | | | iPSYCH | 0.133 | 0.051 | 0.00965 | -0.187 | 0.143 | 0.192 | 0.02 | 0.081 | 0.808 |
| | | | | | Meta | | | | | | | -0.024 | 0.065 | 0.711 |

Chr, chromosome; Beta_SNP, main effect of SNP with corresponding standard error (SE_SNP) and *P* value (P_SNP); Beta_sex, main effect of sex with corresponding standard error (SE_sex) and *P* value (P_sex); Beta_int, interaction effect between SNP and sex with corresponding standard error (SE_int) and *P* value (P_int).

#### Supplementary Table 2

Analysis of alternative genetic models for the seven novel febrile seizures loci in the SSI and iPSYCH discovery cohorts. The table shows effect estimates and corresponding standard errors and P values from an additive genetic model (ADD), a recessive model (REC), a dominant model (DOM), and a full genotype model (AB and BB); *P* values from ANOVA tests of the full genotype model versus the null model, the full model versus the additive model, the full model versus the recessive model, and the full model versus the dominant model. **Table supplied as an additional Excel spreadsheet (online).**

**Supplementary Table 3**

Discovery stage association results for 1843 variants with $P < 1 \times 10^{-4}$ at the seven novel and four known febrile seizures loci. The table is sorted by chromosome and base pair position (GRCh37/hg19) and shows effect and alternative allele; meta-analysis results; SSI discovery cohort results; iPSYCH discovery cohort results; variant annotation and nearest gene; exonic variant class and affected transcripts; SIFT, PolyPhen-2, and FATHMM scores and predictions of missense mutation pathogenicity. Results for the different loci are separated by bold lines. **Table supplied as an additional Excel spreadsheet (online).**

**Supplementary Table 4**

eQTL results based on the BRAINEAC, DICE, eQTL Catalogue, eQTLGen, and GTEx/v8 databases for variants associated with febrile seizures with $P < 1 \times 10^{-4}$ at the seven novel and four known loci. The table is sorted by chromosome and base pair position (GRCh37/hg19) and shows febrile seizures variant, risk allele and discovery *P* value; eQTL database and tissue; Ensembl gene ID, eQTL tested allele, *P* value, effect size, and false discovery rate; febrile seizures risk increasing allele; aligned direction between disease risk and gene expression; variant ID and RefSeq gene name. Results for the different loci are separated by bold lines. **Table supplied as an additional Excel spreadsheet (online).**

**Supplementary Table 5**

GWAS catalog associations ($P < 5 \times 10^{-8}$) for variants associated with febrile seizures with $P < 1 \times 10^{-4}$ at the seven novel and four known loci. The table is sorted by chromosome and base pair position (GRCh37/hg19) and shows febrile seizures variant, risk allele and discovery *P* value; PubMed ID for the relevant GWAS article; trait in the GWAS catalog; SNP and effect allele; type of variant; risk allele frequency; *P* value and accompanying description; odds ratio or beta estimate and 95% confidence interval. Results for the different loci are separated by bold lines. **Table supplied as an additional Excel spreadsheet (online).**

**Supplementary Table 6**

Febrile seizures association analysis of exome sequencing data. For each novel locus, results are shown from gene-based rare-variants association tests for genes within 1 MB of the top SNP at the locus, using the SKAT-O approach. CHROM, BEGIN and END denote chromosomal position on GRCh37/hg19 and NS denotes number of samples. The fraction of individuals with rare variants (FRAC_WITH_RARE), the number of variants (NUM_ALL_VARS), the number of variants after data cleaning (NUM_PASS_VARS) and the number of singleton variants (NUM_SING_VARS) in each gene are given, along with the SKAT-O *P* value and the optimal weight (STATRHO) for the test. Results for each locus is shown in a separate tab. **Table supplied as an additional Excel spreadsheet (online).**

**Supplementary Table 7**

Credible sets of variants with highest posterior probability of being causal at each of the seven novel loci. The sets were found under the assumption of one causal variant at each locus and the posterior probability of one causal variant compared to none is shown as well as the corresponding Bayes factor. In addition minimum, average and median absolute correlation between SNPs in the credible set are shown. The SNPs included in each credible set are listed in order of highest posterior inclusion probability, and the smallest set of SNPs with posterior probabilities summing to 0.95 defines the credible set. Results for each locus is shown in a separate tab. **Table supplied as an additional Excel spreadsheet (online).**

**Supplementary Table 8**

Association results in the iPSYCH and DBDS cohorts for variants at the *IFI44L* and *CD46* loci known to be distinctly associated with febrile seizures occurring after MMR vaccination.

| | | | Alleles | | | | Frequency | | Number | | | |
|---|---|---|---|---|---|---|---|---|---|---|---|---|
| Chr | Position | SNP | Effect | Alt. | Locus | Cohort | Cases | Controls | Cases | Controls | OR (95% CI) | *P* |
| 1 | 79093818 | rs273259 | A | G | *IFI44L* | iPSYCH | 0.715 | 0.699 | 2511 | 46952 | 1.08 (1.01-1.15) | 0.02 |
| | | | | | | DBDS | 0.694 | 0.698 | 922 | 28558 | 0.98 (0.89-1.09) | 0.73 |
| 1 | 208014922 | rs1318653 | T | C | *CD46* | iPSYCH | 0.77 | 0.765 | 2511 | 46952 | 1.03 (0.96-1.10) | 0.44 |
| | | | | | | DBDS | 0.763 | 0.764 | 921 | 28711 | 0.99 (0.89-1.11) | 0.89 |

#### Supplementary Table 9

Association results for the seven novel febrile seizures loci in the SSI discovery cohort taking data about temporal relationship to measles, mumps, and rubella (MMR) vaccination into account.

| Chr | Position (bp) | SNP (effect allele/alternative allele) | Analysis | Effect allele frequency cases | Effect allele frequency controls | Number of cases | Number of controls | OR | *P* value | *P* interaction |
|---|---|---|---|---|---|---|---|---|---|---|
| 1 | 71163715 | rs3001038 (A/G) | MMR+ vs. controls | 0.515 | 0.486 | 925 | 4097 | 1.13 (1.02-1.25) | 0.0185 | |
| | | | MMR+ vs. MMR- | 0.515 | 0.538 | 925 | 1066 | 0.90 (0.79-1.03) | 0.119 | |
| | | | MMR- vs. controls | 0.538 | 0.486 | 1066 | 4097 | 1.23 (1.12-1.36) | 2.35E-05 | |
| | | | All FS vs. controls | 0.527 | 0.486 | 1991 | 4097 | 1.19 (1.10-1.28) | 1.47E-05 | 0.0693 |
| 1 | 206944645 | rs1518111 (T/C) | MMR+ vs. controls | 0.223 | 0.194 | 925 | 4097 | 1.21 (1.07-1.37) | 0.00309 | |
| | | | MMR+ vs. MMR- | 0.223 | 0.208 | 925 | 1066 | 1.09 (0.94-1.28) | 0.252 | |
| | | | MMR- vs. controls | 0.208 | 0.194 | 1066 | 4097 | 1.10 (0.97-1.24) | 0.125 | |
| | | | All FS vs. controls | 0.215 | 0.194 | 1991 | 4097 | 1.15 (1.04-1.26) | 0.00453 | 0.0569 |
| 3 | 49644012 | rs11917431 (T/C) | MMR+ vs. controls | 0.312 | 0.282 | 925 | 4097 | 1.16 (1.04-1.29) | 0.00879 | |
| | | | MMR+ vs. MMR- | 0.312 | 0.305 | 925 | 1066 | 1.04 (0.91-1.19) | 0.557 | |
| | | | MMR- vs. controls | 0.305 | 0.282 | 1066 | 4097 | 1.11 (1.00-1.23) | 0.0446 | |
| | | | All FS vs. controls | 0.308 | 0.282 | 1991 | 4097 | 1.13 (1.04-1.23) | 0.00279 | 0.817 |
| 3 | 56298138 | rs4974130 (C/T) | MMR+ vs. controls | 0.082 | 0.058 | 925 | 4097 | 1.47 (1.21-1.78) | 9.62E-05 | |
| | | | MMR+ vs. MMR- | 0.082 | 0.07 | 925 | 1066 | 1.20 (0.95-1.52) | 0.128 | |
| | | | MMR- vs. controls | 0.07 | 0.058 | 1066 | 4097 | 1.23 (1.01-1.49) | 0.0351 | |
| | | | All FS vs. controls | 0.076 | 0.058 | 1991 | 4097 | 1.34 (1.15-1.55) | 0.000166 | 0.138 |
| 5 | 161508503 | rs1997583 (T/A) | MMR+ vs. controls | 0.105 | 0.092 | 925 | 4097 | 1.16 (0.98-1.37) | 0.0893 | |
| | | | MMR+ vs. MMR- | 0.105 | 0.111 | 925 | 1066 | 0.94 (0.77-1.15) | 0.56 | |
| | | | MMR- vs. controls | 0.111 | 0.092 | 1066 | 4097 | 1.23 (1.06-1.44) | 0.00749 | |
| | | | All FS vs. controls | 0.108 | 0.092 | 1991 | 4097 | 1.20 (1.06-1.36) | 0.00467 | 0.936 |
| 14 | 71323249 | rs244620 (C/T) | MMR+ vs. controls | 0.478 | 0.447 | 925 | 4097 | 1.15 (1.04-1.28) | 0.00899 | |
| | | | MMR+ vs. MMR- | 0.478 | 0.462 | 925 | 1066 | 1.07 (0.94-1.22) | 0.297 | |
| | | | MMR- vs. controls | 0.462 | 0.447 | 1066 | 4097 | 1.06 (0.96-1.18) | 0.222 | |
| | | | All FS vs. controls | 0.469 | 0.447 | 1991 | 4097 | 1.10 (1.02-1.19) | 0.0151 | 0.458 |
| 15 | 64132818 | rs62023078 (G/T) | MMR+ vs. controls | 0.848 | 0.823 | 925 | 4097 | 1.20 (1.05-1.39) | 0.00945 | |
| | | | MMR+ vs. MMR- | 0.848 | 0.849 | 925 | 1066 | 0.99 (0.82-1.18) | 0.875 | |
| | | | MMR- vs. controls | 0.849 | 0.823 | 1066 | 4097 | 1.21 (1.06-1.39) | 0.00422 | |
| | | | All FS vs. controls | 0.848 | 0.823 | 1991 | 4097 | 1.21 (1.09-1.34) | 0.000395 | 0.988 |

Chr, chromosome; bp, base pair; OR, odds ratio (with 95% confidence interval); *P* interaction, *P* value for test of interaction between genotype and vaccination in a model including all febrile seizures cases and controls from the SSI discovery cohort; MMR+, MMR-related febrile seizure cases (febrile seizures occurring in a risk window of 9 to 14 days after the date of MMR vaccination); MMR–, MMR-unrelated febrile seizure cases (febrile seizures occurring 6 weeks or more after vaccination or in an infant with no vaccine exposure); FS, febrile seizures. Controls in the MMR+ vs. MMR-analyses are febrile seizure cases unrelated to MMR vaccination.
